# Repetitive subconcussion results in disrupted neural activity independent of concussion history

**DOI:** 10.1101/2024.03.31.24304818

**Authors:** Kevin Grant Solar, Matthew Ventresca, Rouzbeh Zamyadi, Jing Zhang, Oshin Vartanian, Shawn G Rhind, Benjamin T Dunkley

## Abstract

Concussion is a public health crisis which results in a complex cascade of neurochemical changes in the brain that can have life changing consequences. Subconcussions are considered less serious and were overlooked until recently, but we now realise repetitive subconcussions, such as repetitive head impacts, can lead to serious neurological deficits. Subconcussions are common in contact sports, and the military where certain personnel are exposed to repetitive occupational blast overpressure. Postmortem studies in athletes reveal that cumulative duration of play and force from collisions are better predictors than concussion history for the presence and severity of chronic traumatic encephalopathy – a progressive and fatal neurodegenerative tauopathy, distinct from concussion, thought to be caused by repetitive head impacts, and only diagnosable postmortem – thus, an in vivo predictive biomarker would be game changing. Magnetoencephalography has exceptional temporal sampling for imaging the dynamics of neuronal electrochemical action, and functional MRI shows that functional connectivity is associated with tauopathy patterns. Therefore, both imaging modalities could provide a surrogate biomarker of tauopathy.

In this cross-sectional study, we examined the effects of repetitive subconcussion on neuronal activity and functional connectivity using magnetoencephalography and functional MRI, and on neurological symptoms and mental health in a military sample. For magnetoencephalography and outcome analyses, 81 participants were split into ‘high’ and ‘low’ blast exposure groups using the generalized blast exposure value: *n*=41 high blast (26.4–65.7 years; 4 females); *n*=40 low blast (28.0–63.3 years; 8 females). For fMRI, two high blast male participants without data were excluded: *n*=39 (29.6–65.7 years).

Magnetoencephalography revealed disrupted neuronal activity in participants with a greater history of repetitive subconcussions, including: neural slowing (excessive delta activity) in the right frontal and temporal lobes and subcortical regions (hippocampus, amygdala, caudate, pallidum, thalamus), and functional dysconnectivity in the posterior default mode network (low and high gamma). Notably, these irregularities were independent of concussion or traumatic stress history, and magnetoencephalography revealed functional dysconnectivity not detected with functional MRI. Besides regional slowing and functional disconnection in crucial brain hubs, those with greater blast exposure had poorer neurological outcomes in somatic and cognitive domains, with no blast-related differences in mental health and no associations between neurological symptoms and neuronal activity.

This study suggests that repetitive subconcussions have insidious effects on the brain and that magnetoencephalography provides a potential avenue for both treatment targets by identifying affected brain regions and in prevention by identifying those at risk of cumulative subconcussive neurotrauma.

## 1. Introduction

Concussion, or mild traumatic brain injury (mTBI), involves a complex cascade of neurochemical changes and neurological symptoms that can have life changing consequences.^1^ Repetitive subconcussions were thought to be less serious, and have been largely overlooked up until recently^2–4^ but are now considered insidious, and occur from repetitive head impacts (RHI) common in contact sports^5,6^ and in military settings with occupational repetitive blast overpressure (ReBOP) exposure (e.g., explosions, small arms, heavy weapons).^7–11^ Postmortem studies in athletes reveal that the accumulated duration of play^5,6^ and total force^6^ are better predictors than concussion history for the presence and severity of chronic traumatic encephalopathy (CTE), a progressive and fatal neurodegenerative tauopathy, which is distinct from concussions and persistent postconcussion symptoms (PPCS). Yet, the pathogenic mechanisms of CTE are not well understood,^12^ although histopathology suggests that expression of glial fibrillary acidic proteins at the cortical grey-white matter interface are a neuropathological hallmark of cumulative mild blast or impact neurotrauma.^13^ However, postmortem examinations have shown that CTE was not often detected in the brains of military members, as evidenced by modest neuropathologic changes, and CTE risk ratios were higher for decedents who experienced other traumatic brain injuries (TBI) outside of the military (e.g., contact sports) than those who only had blast exposure or military-related TBI.^14^

Given that CTE and proteinopathic dementias can only be diagnosed postmortem, an in vivo predictive biomarker would be game changing. Positron emission tomography (PET) radiotracer binding to tau with Flortaucipir offers a potential avenue,^15^ with uptake correlating with blast exposure in frontal, temporal, occipital, and cerebellar regions in warfighters and Veterans.^11^ However, a limitation of PET includes its high cost and concern of non-specific binding.^16^ A PET-tau study in Alzheimer’s dementia found that tauopathy colocalized with abnormal synaptic function quantified by magnetoencephalography (MEG),^17^ and another investigation in primary 4-repeat tauopathies showed that tauopathy patterns were associated with functional connectivity quantified by functional magnetic resonance imaging (fMRI).^18^ Together this research suggests that MEG and fMRI can quantify/identify dysfunction associated with tauopathy at a fraction of the cost of PET.

Electrophysiology with MEG offers a highly sensitive, non-invasive means to image the dynamics of neuronal electrochemical action by measuring magnetic fields generated by primary neuronal currents, allowing for micro- to macro-level examinations of neuronal circuits. Neuronal oscillations are important surrogate measures of cell physiology, indexing the rhythmic activity of brain excitability and inhibition, with abnormalities in the temporal pattern of oscillatory activity indicating pathology.^19,20^ MEG can measure neuronal oscillations because of its exceptional temporal sampling rate, whereas fMRI relies on the blood oxygen level dependent (BOLD) signal, a slower haemodynamic proxy for neural activity.

Pathological rhythmic neuronal activity – or ‘*oscillopathies’* – and functional connectivity are important pathogenic markers of psychiatric and neurological disorders, including concussion,^21–26^ post-traumatic stress disorder (PTSD),^21^ Parkinson’s disease (PD),^27^ and Alzheimer’s disease (AD).^28^ “Pathological slowing” of rhythmic activity and changes in functional connectivity are common features of brain injury and neurodegeneration in military personnel and Veterans with concussion.^24^ In neurodegenerative disease specifically, neuronal slowing in PD has been observed in parieto-occipital cortices^27^ and neuronal slowing in AD predicts functional impairment and amyloid burden.^28^

Subconcussions have been studied in the context of RHI, but occupational blast exposure provides a model for the effects of cumulative, repetitive subconcussions common in certain military roles.^29^ Military personnel and Veterans exhibit neurobehavioural symptoms that can mimic persistent post-concussion symptoms, dementia, and PTSD. These include somatic, cognitive, and emotional symptoms and mental health issues (e.g., anxiety, depression).^7–10^ Also, higher lifetime subconcussive blast exposure is associated with poorer brain-related outcomes, including worse neurobehavioural symptoms (e.g., sleep, fatigue).^30^

Although there are functional neuroimaging studies of military personnel who have sustained an mTBI from blast exposure,^31–34^ there are no multimodal studies on the impact of repetitive subconcussions, or those dissociating the effects of concussion on brain function.^35^ There is one MEG study in combat Veterans which identified reduced functional connectivity in blast-related concussion with PTSD^34^; however, they did not disentangle repetitive subconcussive blast exposure from concussion. A magnetic resonance imaging (MRI) study showed that blast exposure resulted in worse cognitive, emotional, and somatic symptoms and smaller cortical volumes in the right superior frontal gyrus.^10^ An fMRI study revealed the effect that a single day of breacher training with acute, multiple blast exposures has on the brain, wherein the researchers gave half of the participants a jugular vein compression collar – those without the collars presented with blast-related impairment in working memory and auditory network connectivity, suggesting that the effects of repetitive subconcussion may be managed by simple protective devices.^36^ In sports, a longitudinal fMRI study showed that high school football players exhibited dysfunctional connectivity across a single season of play despite not showing any symptoms, suggesting pre-symptomatic brain changes.^37^ These studies highlight the value of functional neuroimaging to identify in vivo biomarkers of subconcussive neurotrauma before debilitating symptoms emerge.

The pathological mechanism of repetitive subconcussions – including blast – that might lead to CTE remains largely unknown, but repetitive blast exposure provides a model to investigate pathogenesis and novel markers.^38^ Such a model can inform basic science and provide translatable paths to refine training procedures and improve overall health outcomes for operational readiness.^7–10,30,36,37^ There is evidence that repetitive blast damages the central nervous system and/or the central or peripheral vestibular system due to blast waves entering the ear canal and subsequently causing mechanical reverberations throughout the brain.^39,40^ However, other studies argue that the mechanism extends beyond acceleration–deceleration injuries, due to the effect of blast waves on both air-filled organs and/or organs surrounded by fluid-filled cavities within the body.^41,42^

The purpose of this study was to understand the interaction between lifetime blast exposure estimates, neurobehavioural symptoms, mental health, and brain function outcomes in Canadian Armed Forces (CAF) and Royal Canadian Mounted Police (RCMP) personnel and Veterans exposed to the continuum of repetitive subconcussions from blast overpressure, controlling for concussion and traumatic stress history. The generalized blast exposure value (GBEV) tool^29^ was used to capture blast history – we performed a median split of the cohort into two groups of ‘high’ and ‘low’ blast exposure as described in this other protocol,^43^ to understand the impact and relationships between blast, health outcomes, and neural activity. Multimodal studies suggest that proteinopathic burden and neurodegeneration correlates with neuronal slowing and changes in brain communication^17^ – therefore, we predict that participants with higher blast load will show neuronal slowing (MEG) and abnormal functional connectivity (MEG, fMRI) alongside worse neurobehavioural and mental health symptoms.

## 2. Materials and methods

### 2.1 Participants

Participants were recruited as part of a large, multidimensional, cross-sectional, and longitudinal study with a transdiagnostic design to examine military brain and mental health. Methods of recruitment included dissemination of study information directly to military units, as well as health service channels and philanthropic organizations dealing with military and Veteran brain health (e.g., Project Enlist, Veterans Affairs Canada, and other Veteran’s groups). Participants were eligible for this study if they were active CAF/RCMP personnel or a Veteran of the CAF/RCMP at the time of study enrolment, had sufficient English language skills and cognitive capacity to participate in the study, and had no contradictions to MRI or MEG. Participant data collection occurred from September 2020 to May 2023, and along with all analyses, was conducted at the Hospital for Sick Children in Toronto, ON, Canada.

Initially, 84 total participants were recruited; 3 participants did not complete imaging (MEG or fMRI), and of the 81 remaining, 2 completed the MEG scan but not fMRI. For all MEG and outcome analyses, 81 CAF/RCMP personnel and Veterans from various backgrounds, including those from combat and emergency response roles involving weapons use, armoured, engineering, operators, explosive disposal, and artillery, were divided into two groups based on blast exposure levels assessed by the GBEV questionnaire: *n* = 41 high blast exposure group (aged 26.4 to 65.7 years; mean 48.1 ± 9.0 years; 4 females) and *n* = 40 low blast exposure group (aged 28.0 to 63.3 years; mean 47.8 ± 9.5 years; 8 females). For fMRI analyses, two male participants without fMRI data were removed from the high blast group described above so that: *n* = 39 high blast group (aged 29.6 to 65.7 years; mean 48.6 ± 8.5 years; 4 females). The GBEV is a tool for standardizing a lifetime of blast exposure.^29^ It quantifies units of blast exposure by querying participants about exposure over a lifetime across five categories of blast type/severity. This study was approved by the Hospital for Sick Children Ethics Board. Written informed consent was obtained from all participants in accordance with the Declaration of Helsinki.

### 2.2 Neurobehavioural and psychiatric outcomes

All participants completed neurobehavioural and mental health assessments which included: the Generalized Anxiety Disorder 7 screener (GAD7)^44^; Patient Health Questionnaire (PHQ9)^45^; PTSD Checklist Military Version (PCL-M)^46^; and the neurobehavioural symptom evaluation from the Sports Concussion Assessment Tool 5 (SCAT5).^47^ Participants also completed brief screeners related to diagnosed lifetime concussion history, using the Acute Concussion Evaluation (ACE)^48^ and defined by Department of Defence/Veterans Affairs guidelines for mTBI as injury resulting in loss of consciousness <30 minutes, post traumatic amnesia <24 hours, and a Glasgow Coma Score of 13 or more. Traumatic stress history was also estimated with the Brief Trauma Questionnaire (BTQ).^49^

### 2.3 Magnetoencephalography acquisition and analysis

Resting state MEG data were acquired on a 151 channel CTF system at the Hospital for Sick Children in Toronto. Participants were scanned in the supine position with fiducial coils attached at the nasion and bilateral preauricular pits for head motion recording. Data sampling rate was 600 Hz, bandpass filtered offline (high-pass, 1 Hz; low-pass, 150 Hz; notch, 60 Hz), and analyzed in the Fieldtrip Toolbox.^50^ Sensor level time series were mean centred. Cardiac and ocular artefacts were removed using independent component analysis (ICA). Time-series data were divided into 10 s epochs and the maximum number were retained from the 5 minute recording based on the following exclusion criteria: (a) head position did deviate more than 5 mm during a given epoch; (b) epochs that contained superconducting quantum interference device resets or exceeded a ±2 pT threshold following ICA component rejection.

MEG co-registration was performed using a single-shell head model per participant based on their anatomical T1-weighted MRI data normalized to Montreal Neurological Institute (MNI) space. A beamformer was used to resolve time series data from 90 regions of the Automated Anatomical Labelling (AAL) atlas^51^ with the parcel centroid of each AAL region used as the node location for beamformer reconstruction for which the Fieldtrip^50^ linearly constrained minimum variance vector beamformer^52^ was used, with 5% Tikhonov regularization. At each AAL node, lead fields were calculated from the template single shell head model for a unit current dipole across three dimensions. Beamformer weights of nodal neural activity were calculated through projection of the sensor weights along the dimensional axis with the highest singular value decomposition variance which produces a neural activity time series for each node.

The retained time series data from virtual electrodes were z-scored (i.e., mean centred, variance normalised). These broadband regional time courses per node were then filtered into delta (1-3 Hz), theta (3-7 Hz), alpha (8-14 Hz), beta (15-25 Hz), low gamma 1 (30-55 Hz), low gamma 2 (65-80 Hz), and high gamma (80-150 Hz) ranges. To reduce artificially inflated coupling that can result from beamformer leakage, a symmetric orthogonalization correction procedure was used.^53^ Welch’s Method was used to obtain the power spectrum density (PSD) at each AAL node. Lobe-wise average power was calculated per hemisphere for each participant by averaging across each AAL node within a given lobe.^54^ The Hilbert transform was applied to the band-limited time course to generate instantaneous estimates of the amplitude envelope, which was then down-sampled to 1 Hz.^55^ Pearson correlations were calculated between all AAL node pairs to quantify functional coupling. Amplitude envelope coupling (AEC) was chosen over other measures of neural communication (for example, phase synchronisation) as it is the most reliable measure of connectivity across sessions and over individuals,^21,56,57^ pointing to the greatest replicability as well as the lowest susceptibility to co-registration related errors.^58^

### 2.4 MRI acquisition and analysis

MRI data were acquired on a 3T Siemens PrismaFit scanner with a 20-channel head and neck coil at the Hospital for Sick Children in Toronto. For source localization of MEG and fMRI data, 3D magnetization-prepared rapid gradient echo T1-weighted images were acquired with 0.8 mm isotropic voxel size in 5:01 minutes (repetition time, TR, 1870 ms; echo time, TE, 3.1 ms; inversion time, TI, 945 ms; field-of-view, FOV, 240×256 mm with 240 0.8 mm slices).

Resting state fMRI data were acquired with 3 mm isotropic voxel size in 5 minutes (TR, 1500 ms; TE, 30 ms) and standard preprocessing was completed with the Data Processing Assistant for Resting-State fMRI toolbox^59^ and the SPM8 toolbox^60^ (http://www.fil.ion.ucl.ac.uk/spm). First, slice time correction was applied to the interleaved sequence acquisition, then realignment of the volumes was accomplished by calculating a six-parameter rigid body spatial transformation.^61^ None of the participants exhibited >3 degrees rotation or >3 mm displacement in any direction across the acquisition; therefore, no volumes were discarded. Next, normalization to MNI space was performed through unified T1-weighted image segmentation. The global signal was filtered out along with nuisance covariates including head motion parameters, white matter (WM) signal, and cerebrospinal fluid (CSF) signal to minimize the effects of non-neuronal oscillations and motion.^62–65^ Next, images were band-pass filtered at 0.01-0.1 Hz to retain low frequency oscillations in the resting state fMRI which are believed to represent neuronal activity,^66^ while suppressing low frequency drifts and high frequency noise resulting from other physiological activity.^67,68^ BOLD time domain signals were extracted from every voxel of the same 90 AAL atlas regions as MEG.^51^ For each individual AAL region, the mean timeseries were calculated across a parcel, then used to estimate functional connectivity with Pearson correlations.^69^

### 2.5 Statistical analysis

Statistical analyses were performed using MATLAB version R2022b, JASP version 0.18.0 (JASP, 2023), and the Network-Based Statistic (NBS) toolbox.^70^ False positives due to multiple comparisons were controlled using the Benjamini-Hochberg false discovery rate (FDR-BH) method at a significance threshold of *p* < 0.05. There are known effects of sex, age, concussion, and psychological trauma in these brain and outcome measures; thus, these confounders were controlled for as described below.

#### 2.5.1 Participant demographics and outcomes

Analyses of covariance (ANCOVA) were used to test for blast-related group differences in outcome measures (GAD7, PHD9, PCL-M, SCAT5) while controlling for age, sex, traumatic stress history (BTQ), and number of diagnosed concussions (ACE). ANCOVA was also used to test for group differences in outcomes, stratified by concussion/no concussion history, while controlling for age and sex.

#### 2.5.2 Power spectrum density analysis

ANCOVAs were used to test for blast-related group differences in power across all seven frequency bands across hemispheric lobes by calculating the average power for each AAL atlas node in the left and right frontal, temporal, parietal, and occipital lobes, and subcortical regions (grouping hippocampus, amygdala, caudate, pallidum, thalamus) while controlling for age, sex, psychological trauma, and number of concussions.

#### 2.5.3 Network based analysis

The NBS toolbox was used to test group differences in functional connectivity within eight networks based on AAL atlas nodes (anterior DMN, aDMN; posterior DMN, pDMN; sensorimotor network, SMN; salience network, SN; memory network, MN; visual network, VN; central executive network, CEN; attention network, AN) in the seven MEG frequency bands and single fMRI connectomes, while controlling for age, sex, psychological trauma, and number of concussions (statistical model: threshold = 3.1; F-test; 5000 permutations; NBS method; Extent component size).

#### 2.5.4 Concussion group analysis

Conveniently, of the 81 total participants, *n* = 38 (aged 26.4 to 65.7 years; mean 48.6 ± 9.5 years; 5 females) had a history of a clinical and confirmed diagnosis of at least one concussion, and the remaining *n* = 43 (aged 28.0 to 61.9 years; mean 47.3 ± 8.9 years; 7 females) did not; therefore, we also conducted an exploratory between groups analysis based on concussion history, covarying for age, sex, psychological trauma, and blast exposure as quantified by the GBEV, to examine concussion specific changes in brain function with MEG. For the fMRI analyses, the two male participants without fMRI data were removed: one from the concussion group (*n =* 37; aged 29.6 to 65.7 years; mean 49.2 ± 8.9 years; 5 females) and another from the non-concussion group (*n =* 42; aged 28.0 to 61.9 years; mean 47.3 ± 9.0 years; 7 females).

#### 2.5.5 Brain function and outcome association analyses

As a post-hoc analysis, regressions were conducted between any brain measures (MEG power or connectivity, fMRI connectivity) and outcomes (mental health, neurological) that showed significant blast-level group differences, while controlling for age, sex, psychological trauma, and number of concussions.

## 3. Results

### 3.1 Demographics

The age of the high blast group (*n* = 41; range: 26.4 to 65.7 years; mean: 48.1 ± 9.0 years) did not differ from the low blast group (*n* = 40; range: 28.0 to 63.3 years; mean: 47.8 ± 9.5 years), nor did sex ratios (high blast group: 37 males, 4 females; low blast group: 32 males, 8 females) (**Figure 1A**), and there were no age or sex differences when the two high blast male participants were removed for fMRI analyses due to lack of data (*n* = 39; range: 29.6 to 65.7 years; mean 48.6 ± 8.5 years; 35 males, 4 females). Additionally, when re-stratifying the cohort by concussion history, the age of the concussion group (*n* = 38; range: 26.4 to 65.7 years; mean: 48.6 ± 9.5 years) did not differ from the non-concussion group (*n* = 43; range: 28.0 to 61.9 years; mean: 47.3 ± 8.9 years), nor did sex ratios (concussion group: 33 males, 5 females; non-concussion group: 36 males, 7 females), and there were no age or sex differences when the two male participants without fMRI data were removed, including one from the concussion group (*n* = 37; range: 29.6 to 65.7 years; mean: 49.2 ± 8.9 years; 32 males, 5 females) and another from the non-concussion group (*n* = 42; range: 28.0 to 61.9 years; mean: 47.3 ± 9.0 years; 35 males, 7 females) (**Figure 1B**).

**Figure 1:**
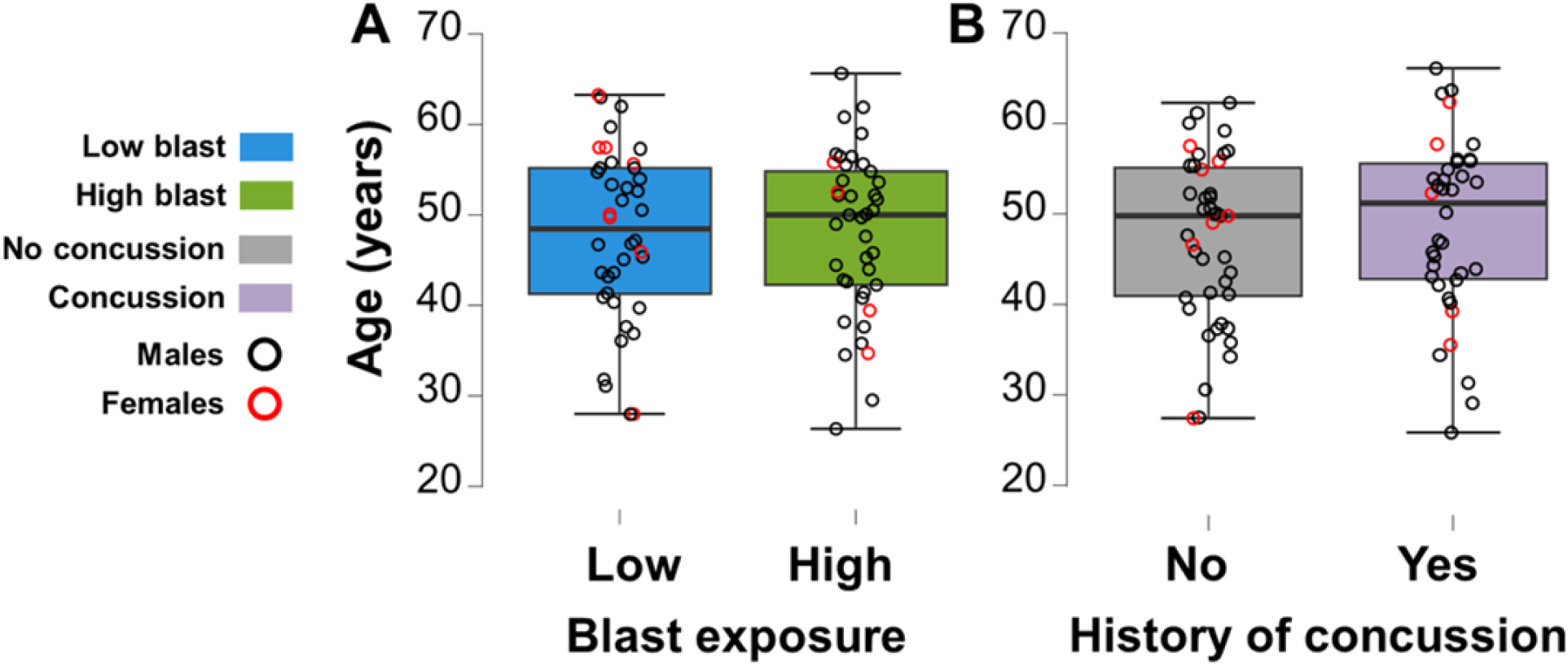
Demographics. Age distribution for the entire cohort divided by (**A**) lifetime blast exposure and (**B**) concussion history.

### 3.2 Neurological symptoms due to repetitive subconcussive overpressure, not concussion history

The high blast exposure group showed significantly worse overall neurobehavioural symptom severity (**Figure 2A**), driven by poorer outcomes on somatic (**Figure 2B**) and cognitive subscales (**Figure 2C**), with no blast-related differences in emotional (**Figure 2D**) or sleep symptoms (**Figure 2E**) when controlling for age, sex, psychological trauma history, and number of diagnosed concussions.

**Figure 2:**
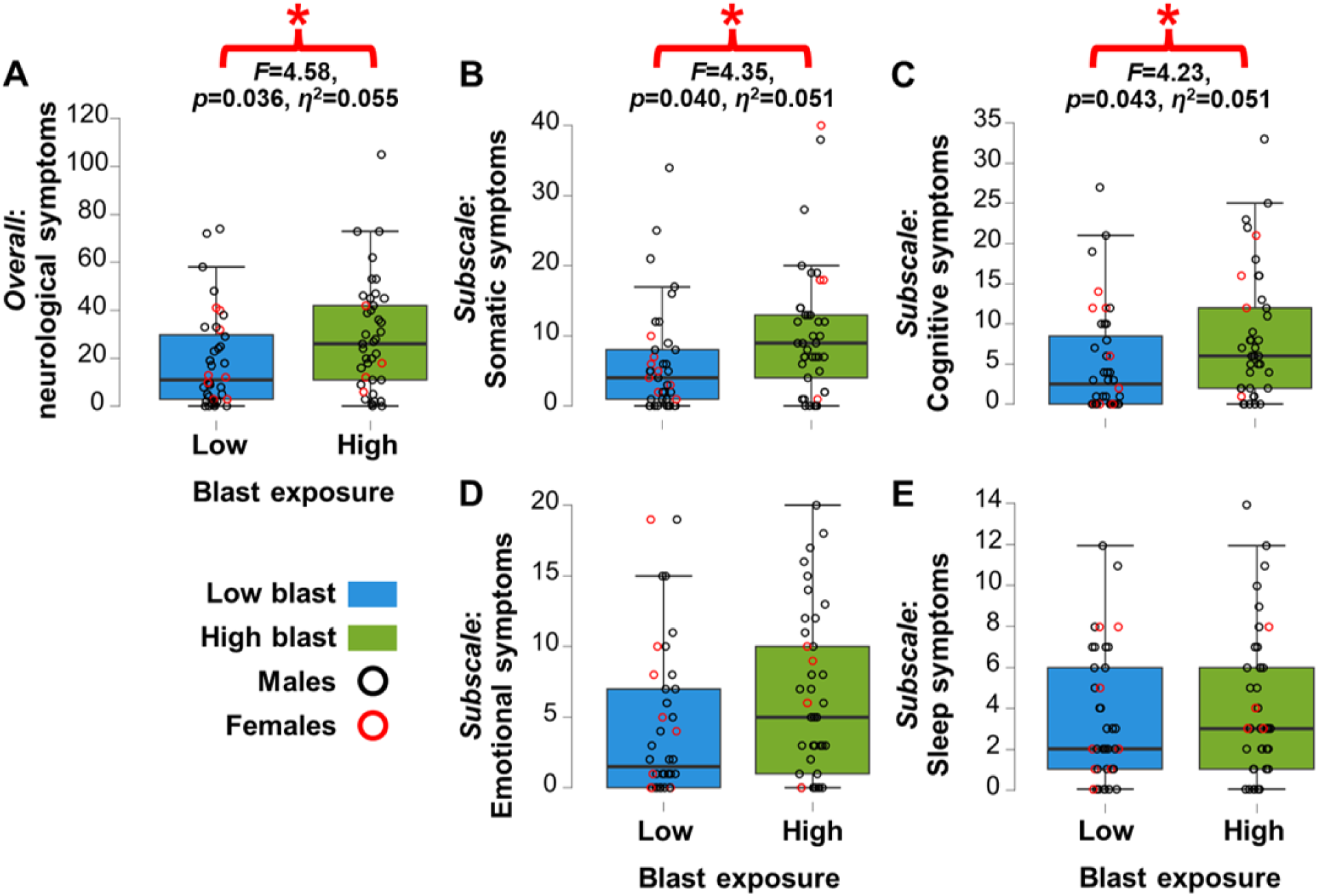
Neurobehavioural outcomes are worse in those with greater blast exposure, independent of concussions or traumatic stress history. (**A**) Overall neurological symptom severity was significantly higher in the high blast exposure group (green) than the low blast group (blue) (*F*(1, 75) = 4.58, *p* = 0.036, *η*^2^ = 0.055), driven by significantly worse (**B**) somatic (*F*(1, 75) = 4.35, *p* = 0.040, *η*^2^ = 0.051) and (**C**) cognitive symptoms (*F*(1, 75) = 4.23, *p* = 0.043, *η*^2^ = 0.051) – with no significant group differences in the subscale measures of (**D**) emotional or (**E**) sleep symptoms, when controlling for age, sex, number of diagnosed concussions, and psychological trauma (unadjusted estimates: overall neurological symptom severity, *F*(1, 79) = 5.00, *p* = 0.03; somatic symptoms, *F*(1, 79) = 4.34, *p* = 0.04; cognitive symptoms, *F*(1, 79) = 3.72, *p* = 0.06).

There were no group differences between high and low blast in the number of diagnosed concussions (**Figure 3A**), psychological trauma severity (**Figure 3B**), and mental health outcomes, including depression (**Figure 3C**), anxiety (**Figure 3D**), or PTSD (**Figure 3E**) symptom severity. Moreover, there were no significant group differences for neurobehavioural, anxiety, depression, or PTSD symptoms in the concussion stratified analysis when controlling for age, sex, psychological trauma, and blast exposure history. These results suggest that the elevated neurological symptoms experienced by individuals with greater blast exposure is due to repetitive subconcussions and not concussion or traumatic stress history.

**Figure 3:**
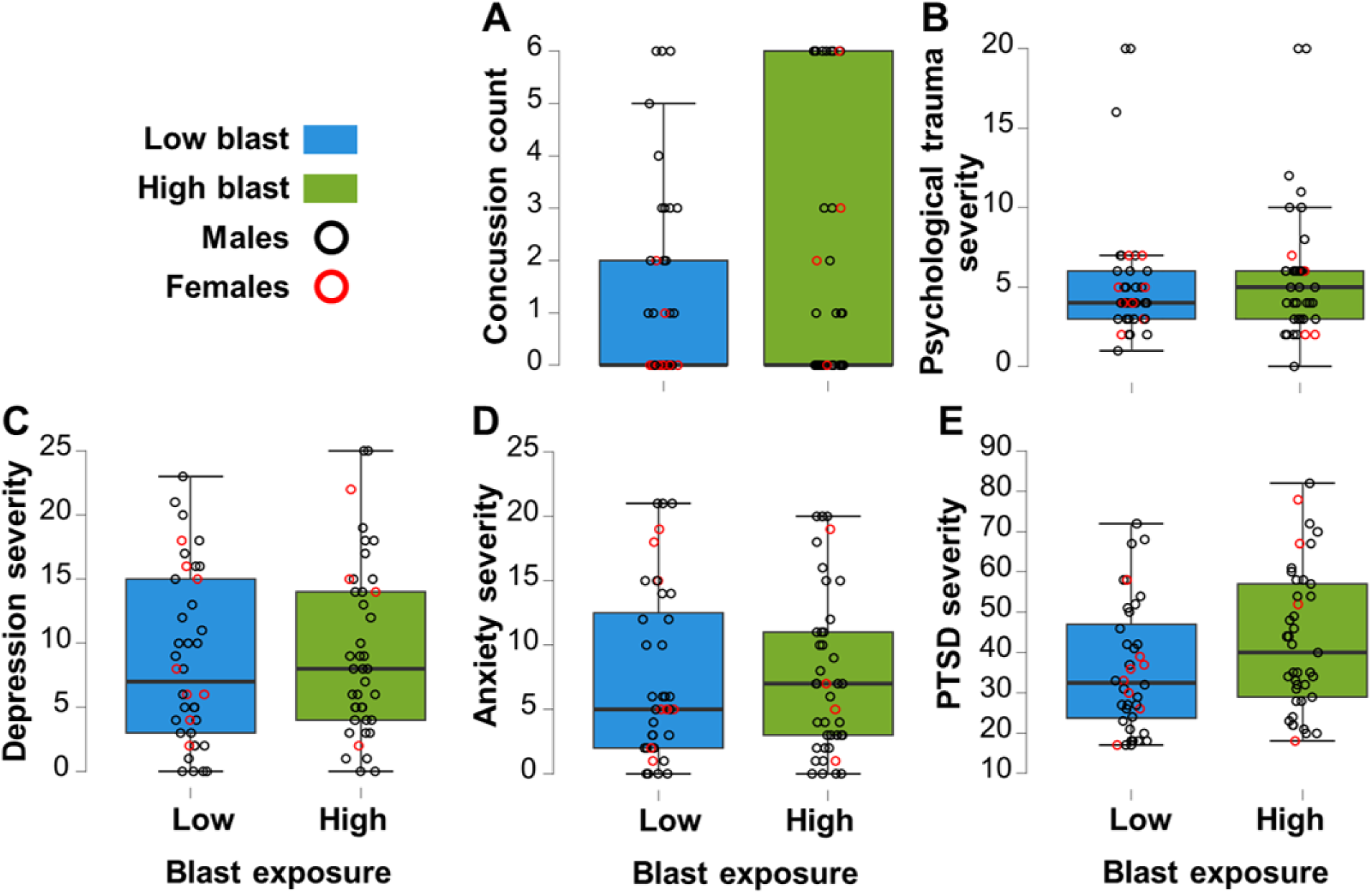
Concussion history and mental health outcomes did not differ as a function of blast exposure. There were no significant differences between the high blast exposure group (green) and the low blast group (blue) in (**A**) the number of diagnosed concussions, (**B**) psychological trauma index (BTQ), nor in mental health outcomes, including (**C**) depression (PHQ9), (**D**) anxiety (GAD7), and (**E**) PTSD (PCL-M) severity.

### 3.3 Neural slowing is evident in fronto-temporal and subcortical regions after high blast exposure

Comparisons of lobe-wise power spectrum reveal larger alpha amplitude in the high blast group, but otherwise comparable and expected 1/f power spectrum curves for both groups (**Figure 4**).

**Figure 4:**
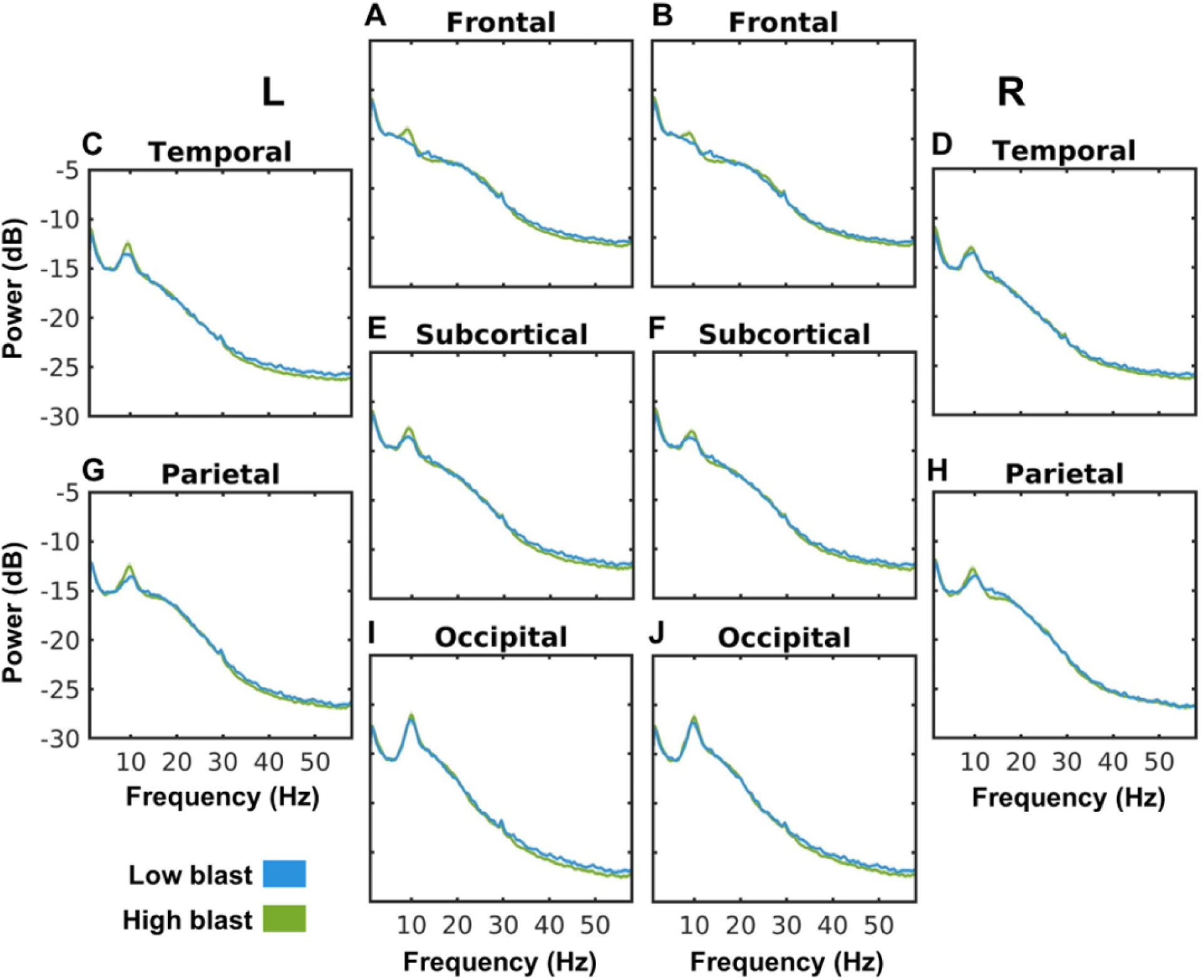
Lobe-wise power spectrum as a function of blast exposure. (**A-J**) Lobe-wise power spectrum for low (blue) and high (green) blast exposure groups for 1-30 Hz range. (**E**, **F**) Subcortical regions include hippocampus, amygdala, caudate, pallidum, and thalamus.

Delta activity (1-3 Hz) was significantly higher in the right frontal and right temporal lobes, as well as in right subcortical regions (hippocampus, amygdala, caudate, pallidum, thalamus) in the high blast compared to low blast group when controlling for age, sex, traumatic stress, and number of diagnosed concussions (**Figure 5**), but not for any other lobes at the delta frequency. There were no group differences for any other of the six frequency bands.

**Figure 5:**
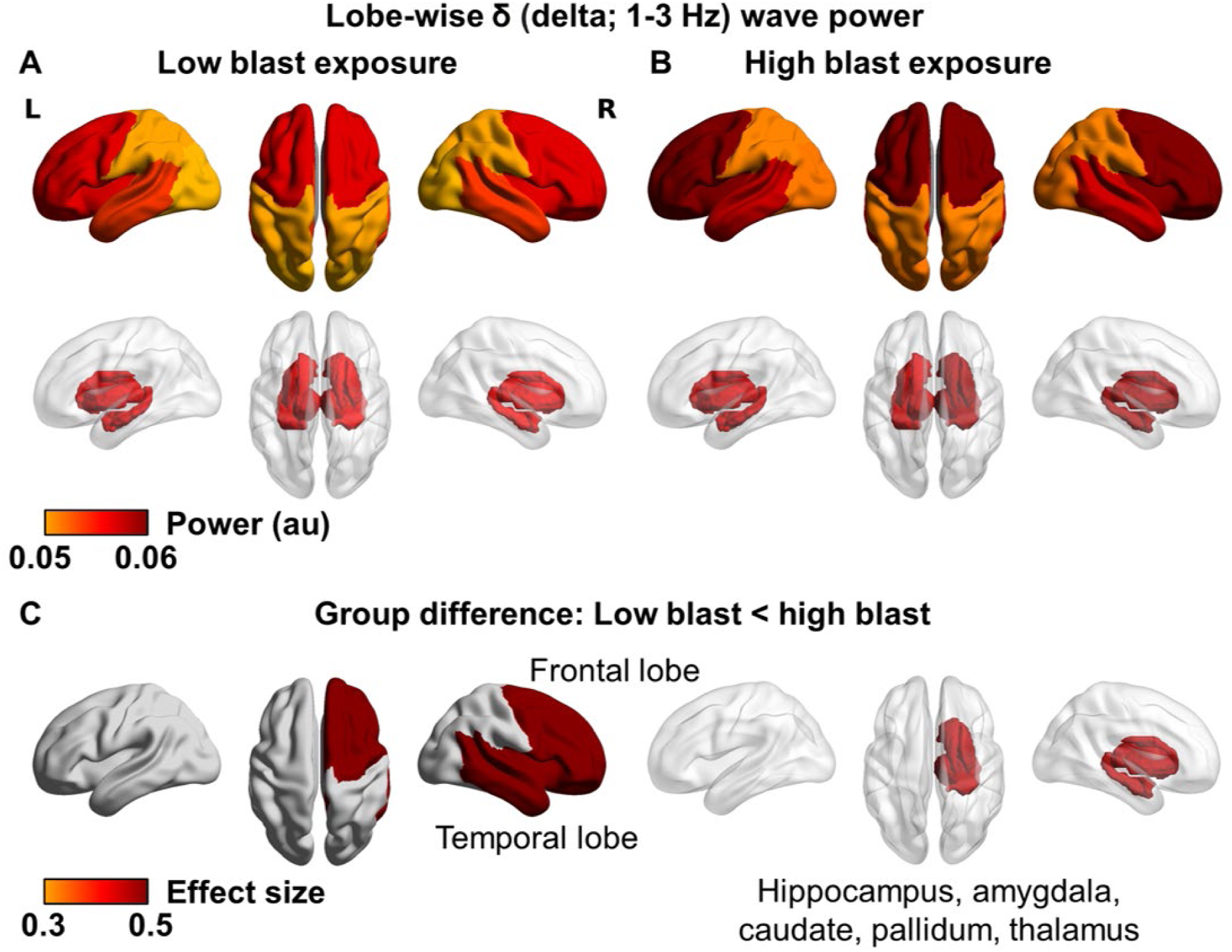
Repetitive subconcussions cause neural slowing in fronto-temporal and subcortical regions, independent of concussions or traumatic stress history. Lobe-wise delta (1-3 Hz) activity in the (**A**) low and (**B**) high blast exposure groups, with (**C**) the high blast exposure group showing significantly higher delta power in the right frontal (*F*(1, 75) = 8.85, *p* = 0.004, *η*^2^ = 0.10) and temporal (*F*(1, 75) = 7.95, *p* = 0.006, *η*^2^ = 0.092) lobes and subcortical regions (hippocampus, amygdala, caudate, pallidum, thalamus) (*F*(1, 75) = 7.27, *p* = 0.009, *η*^2^ = 0.084) (unadjusted estimates: right frontal lobe, *F*(1, 79) = 1.76, *p* = 0.19; right temporal lobe, *F*(1, 79) = 1.34, *p* = 0.25; right subcortical regions, *F*(1, 79) = 1.04, *p* = 0.31).

Moreover, there were no group differences when comparing diagnosed concussion versus no concussion history in regional power when controlling for age, sex, trauma, and blast exposure. Together, these results suggest that neural slowing in the right fronto-temporal lobes and subcortical regions is due to repetitive subconcussive neurotrauma and not concussion history.

### 3.4 Decreased pDMN connectivity in high blast exposure as measured by MEG but not fMRI

MEG revealed significant reductions in pDMN functional connectivity for the high blast group at low gamma (30-55 Hz; 7 nodes and 7 edges; **Figure 6A**) and high gamma (80-150 Hz; 14 nodes and 17 edges; **Figure 6B**).

**Figure 6:**
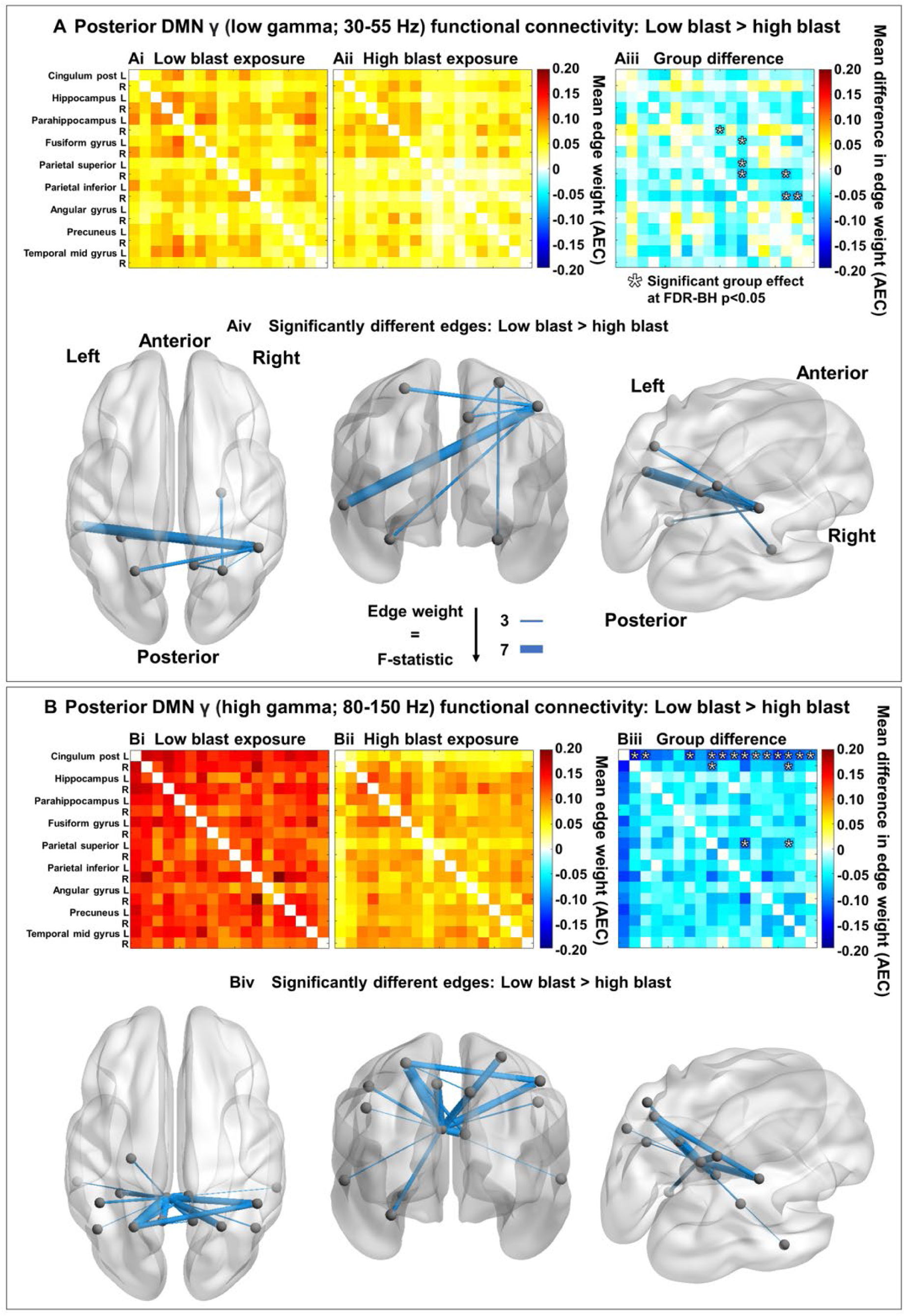
Repetitive subconcussions cause posterior default mode network dysconnectivity, independent of concussions or traumatic stress history. (**A**) MEG low gamma (30-55 Hz) range and (**B**) high gamma (80-150 Hz) range from the 18 total nodes of the pDMN with (**Ai**, **Bi**) low and (**Aii**, **Bii**) high blast exposure group connectivity matrices scaled by mean edge weight (AEC) with significantly lower connectivity in the high blast relative to the low blast exposure group for 7 nodes and 7 edges at low gamma (**Aiii**, **Aiv**; *F*(1, 75) = 4.20, *p* = 0.044, *η*^2^ = 0.053) and 14 nodes and 17 edges at high gamma (**Biii**, **Biv**; *F*(1, 75) = 5.31, *p* = 0.024, *η*^2^ = 0.066). Warm elements in the (**Ai**, **Aii**, **Bi**, **Bii**) group mean matrices indicate higher connectivity and cool colours indicate lower connectivity; warm elements in the (**Aiii**, **Biii**) group difference matrices (calculated from the low blast group means, **Ai** and **Bi**, subtracted from the high blast group means, **Aii** and **Bii**, respectively) indicate higher connectivity in the high blast group and cool colours indicate lower connectivity in the high blast group. At low gamma, the node with the highest degree was the right inferior parietal (**Aiii**, **Aiv**) and at high gamma, it was the left posterior cingulate (**Biii**, **Biv**) (unadjusted estimates: pDMN low gamma, *F*(1, 79) = 2.47, *p* = 0.12; pDMN high gamma, *F*(1, 79) = 3.26, *p* = 0.075).

For low gamma connectivity, the high blast group exhibited dysconnectivity in the pDMN including the right parahippocampus, left fusiform gyrus, bilateral superior parietal cortex, right inferior parietal cortex, right precuneus, and left middle temporal cortex (marked with asterisks, **Figure 6A**) – with no differences in the other seven networks (**Supplemental Figures S1, S2**). For high gamma connectivity, the high blast group exhibited dysconnectivity across pDMN areas including the bilateral posterior cingulate cortex (PCC), left hippocampus, left fusiform gyrus, bilateral superior and inferior parietal cortex, bilateral angular gyrus, bilateral precuneus, and bilateral middle temporal cortex (marked with asterisks, **Figures 6B**) – with no differences in the other seven networks (**Supplemental Figures S3, S4**).

The regions with the greatest difference (edges scaled by the F test statistic, **Figures 6A** & **6B**) were mainly interhemispheric connections across both gamma ranges in the pDMN. There were no blast exposure group differences in any of the other individual networks at any frequency band, including the AN, CEN, aDMN, MN, SMN, SN, and VN. In contrast to the MEG results, fMRI did not reveal any significant blast-related differences (**Supplemental Figures S5, S6**). Additionally, there were no significant group differences in either MEG or fMRI connectivity when stratified by concussion history. These results suggest that (1) the dysconnectivity of the pDMN in the high blast group is due to repetitive subconcussions, and not concussion history; (2) MEG is more sensitive to network dysregulation from repetitive subconcussions than fMRI.

### 3.5 MEG delta power and functional dysconnectivity does not correlate with neurobehavioural outcomes

Despite group differences in delta activity (right frontal and temporal lobes and subcortical regions), pDMN functional connectivity, and neurological symptom outcomes (overall neurological symptom severity, and subscales of cognitive and somatic symptom severity), there were no significant associations between MEG delta power or functional connectivity and neurological symptom severity when controlling for age, sex, psychological trauma, and number of concussions (**Figure 7A-I**). For functional connectivity in the pDMN, node strength per participant was calculated for the node with the highest degree of connections with blast-related differences – namely, the right inferior parietal hub at low gamma and the left posterior cingulate hub at high gamma. There were also no significant associations between MEG functional connectivity and neurological symptom severity when controlling for age, sex, psychological trauma, and number of concussions (**Figure 7J-O**).

**Figure 7:**
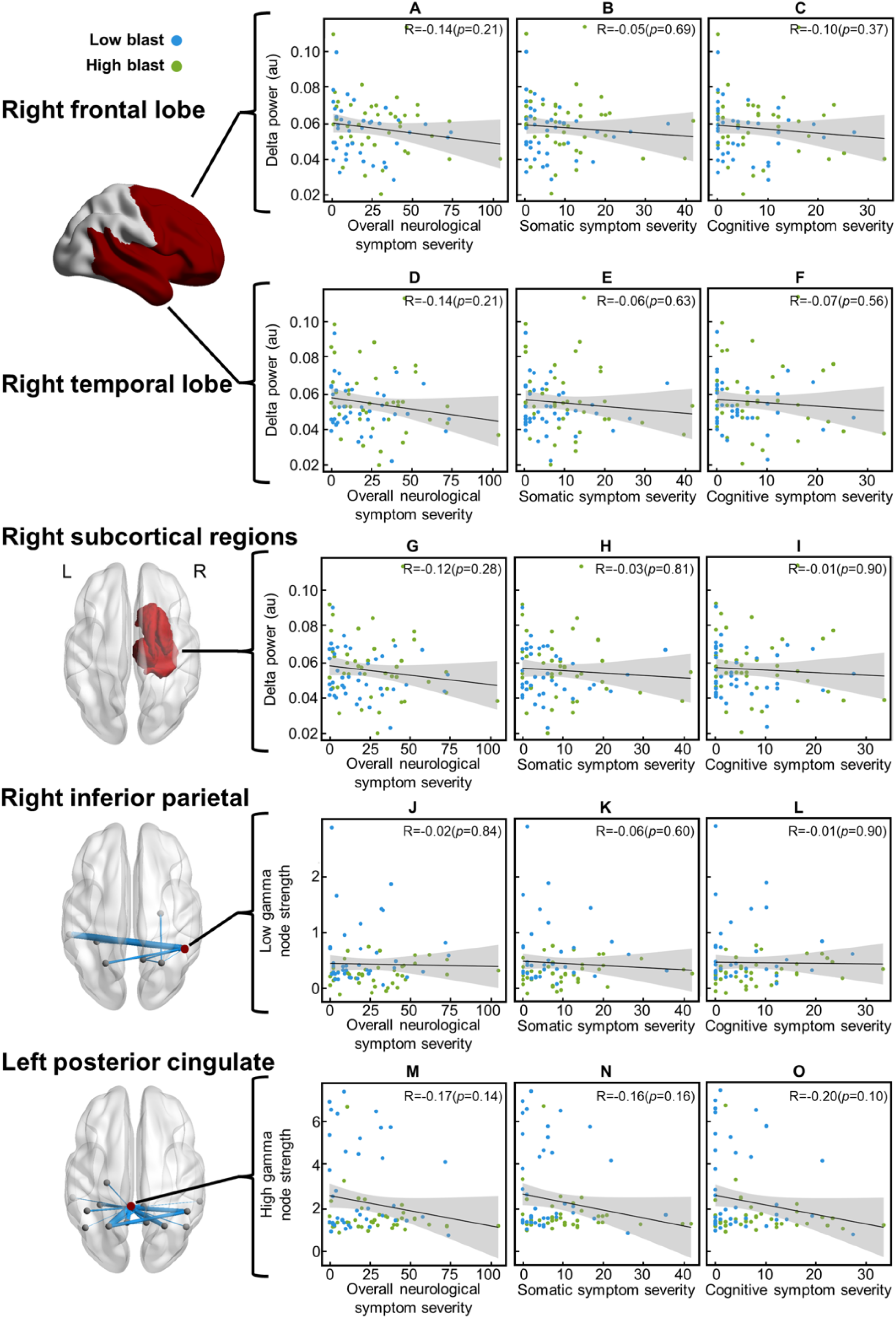
No blast-related associations between neuronal slowing and dysconnectivity with symptoms. There were no significant associations between (**A-I**) neuronal slowing or (**J-O**) dysconnectivity and symptoms; participants with low blast exposure are indicated in blue and those with high blast exposure in green (shaded area represents 95% confidence interval). (**A-C**) Right frontal lobe delta power, (**D-F**) right temporal lobe delta power, (**G-I**) right subcortical regional (hippocampus, amygdala, caudate, pallidum, thalamus) delta power, (**J-L**) right inferior parietal cortex low gamma pDMN connectivity node strength, and (**M-O**) left posterior cingulate cortex high gamma pDMN connectivity node strength did not significantly correlate with overall neurological symptom severity, somatic symptom severity, or cognitive symptom severity.

## 4. Discussion

### 4.1 Summary

We present the first multimodal functional neuroimaging evidence of dysregulated neurophysiological functioning in a sample of military members and Veterans with a history of repetitive subconcussions, resulting from varying degrees of exposure to blast overpressure – importantly, these results were observed independently of concussion or traumatic stress history. Along with regional slowing of activity, indexed by increased delta activity, and functional dysconnectivity in key brain hubs, those with higher blast exposure also exhibited worse neurological symptoms, especially in the somatic and cognitive domains, with no blast-related differences in mental health outcomes. Despite regional slowing and dysconnectivity and worse neurological symptoms in higher blast exposure, there were no significant associations between MEG power and network functional connectivity and neurological symptom severity.

### 4.2 Neural slowing in fronto-temporal and subcortical regions

Those with greater blast exposure exhibited neural slowing – elevated delta activity – in fronto-temporal and subcortical regions that was independent of concussion history. Moreover, we observed functional dysconnectivity in the pDMN – an effect that was absent in our fMRI measurements using the exact same cohort. Despite group blast-related differences in neurological outcomes and neuronal activity and functional connectivity, there were no associations between neurological outcomes and functional measures.

Animal models show a dose-response curve of blast exposure frequency with tauopathy^71^ that results in abnormal neuronal activity and neurobehavioural deficits.^71–78^ This work is consistent with our findings which show worse cognitive, somatic, and physical symptoms, neural slowing in the right fronto-temporal and subcortical regions, and lower functional connectivity in the pDMN in those with greater subconcussive load. Rodent studies have shown that even a single blast (i.e., mild-moderate shockwave) can have insidious effects, resulting in poorer memory function and elevated markers of neurodegeneration, microglia activation, and gliosis with decreased mature neurons in the prefrontal cortex and limbic areas, including the hippocampus, at three months post-exposure – suggesting these areas are particularly susceptible to the effects of blast.^78^

Another rodent study reported brain-wide and hippocampal CTE-linked neuropathology, including myelinated axonopathy, chronic neuroinflammation, and neurodegeneration two weeks following a single blast, and at one month, cognitive impairment, reduced synaptic plasticity, and slowed axonal conduction velocity^72^ – mechanistically, this would explain our observations of neural slowing. Importantly, the neurobiological mechanism of neuronal slowing differs between acute brain injury and neurodegenerative disease processes: slowing in brain injury is putatively caused by axonal shearing at the grey-white matter boundary,^79^ whereas in neurodegeneration, slowing is caused by proteinopathy, such as microtubule degradation, hyperphosphorylation of tau, and resultant neurofibrillary tangles (NFT).^80,81^ For example, in both civilian and military mTBI, multimodal neuroimaging with MEG detected neuronal slowing and diffusion tensor imaging (DTI) found proximal axonal / white matter damage.^82^

Our data suggest that delta oscillations, a stable in vivo biomarker of neural slowing, could have clinical utility in a broad range of neurodegenerative diseases, including Parkinson’s disease (PD),^27^ Alzheimer’s disease (AD),^28^ and other dementias such as fronto-temporal and vascular dementia.^83^ A study by Goldstein *et al*.^72^ reported CTE-linked cortical and hippocampal neuropathology; it included a complementary ex vivo analysis of brains from blast-exposed military veterans, American football athletes, and typical controls which revealed CTE-linked tauopathy as identified by tau-immunoreactive NFTs in the frontal, parietal, and temporal cortices.^72^ Together, these findings suggest that blast exposure causes neurodegeneration which leads to abnormal neuronal activity, providing a potentially viable non-invasive biomarker of disease states.

### 4.3 High-frequency mediated DMN dysconnectivity

We observed high frequency pDMN dysconnectivity, converging on the PCC. The PCC is well-established as a multimodal, polysensory, rich club node, with a metabolic rate on average ∼40% higher than the rest of the brain.^84^ Prior studies have shown PCC dysfunction in numerous acquired brain injury and neurodegenerative states, with PCC lesions severely impacting dynamical functional network configurations associated with general cognitive performance.^85,86^ Patients with mTBI have shown reduced DMN connectivity involving the posterior cingulate,^87^ and interhemispheric posterior DMN functional connectivity is predictive of the level of consciousness in TBI.^88^ Moreover, a longitudinal pre-/post-season fMRI study by DeSimone *et al*.^89^ reported that the number of subconcussive RHIs were key in causing reduced DMN functional connectivity (which was not present pre-season) in a group of football players in the absence of concussion after only a single season of play relative to control athletes from non-contact sports, particularly including dysconnectivity with the left PCC. In our study, we found reduced posterior DMN functional connectivity in the gamma band, gamma being a mechanism thought to selectively and flexibly couple proximal or distant cortical regions to temporally organize neuronal activity through the action of interneurons.^90^

Animal models of tauopathy suggest that tau protein propagation in the brain is promoted by neuronal activity.^91–93^ A recent study of patients with participants along the AD spectrum (preclinical, mild cognitive impairment, and dementia) indicated that tau (identified with PET) spread along functionally connected (identified by MEG) brain regions.^94^ Another AD study in amyloid-positive patients showed lower functional connectivity (identified by fMRI) in the DMN for those with elevated tau (identified with PET) but higher functional connectivity for those with lower tau.^95^ Along this line of work, a study by García-Colomo *et al*.^96^ reported longitudinal increases in functional connectivity (identified by MEG) in correlation with tau (identified with PET) in the precuneus in individuals with a family history of AD relative to those without – a brain region which was also identified here as part of the disconnected pDMN. This work suggests that changes in functional connectivity as identified by MEG can be indicative of tauopathy, in turn allowing us to not only track disease progression but also to pre-emptively predict where tauopathy will spread based on functional connectivity. MEG could be utilized in the treatment and diagnosis of brain injury by identifying regions of the brain in affected individuals to deliver precision treatment to reduce further spreading and predict individuals at risk and intervene before tauopathy spreads.

### 4.4 Association with symptoms

Although worse self-reported neurological symptoms were present with greater blast exposure, there were no significant direct associations between neuronal slowing or dysconnectivity and symptoms. However, neuronal slowing in fronto-temporal and subcortical regions is likely pathogenic in terms of neurobehavioural deficits. The frontal lobe is involved in executive functioning including mental flexibility, goal-directed behaviours, language, learning, and memory, and related processes like attention.^97^ The hippocampus – part of our subcortical analysis – is involved in multiple aspects of memory, including visual and auditory working and episodic memory,^98–101^ as well as a breadth of other processes including executive functioning, attention, social behaviours, spatial navigation, and language.^101,102^ Together, the self-reported cognitive difficulties align with the regional dysfunction we observed. We believe that additional research is needed to explore more fully the association between MEG network functional connectivity and neurological symptom severity, perhaps based on a more comprehensive set of outcome measures known to be impacted by ReBOP.

### 4.5 Translational use

Notably, MEG but not fMRI revealed pDMN functional dysconnectivity in greater blast exposure, suggesting increased sensitivity to detect network-level effects – these effects were in the gamma range, high-frequency oscillatory activity to which fMRI is blind. Gamma activity reflects the coordinated action of neural excitability and inhibition, driven by local microcircuits involving excitatory pyramidal neurons and inhibitory interneurons, described as the pyramidal-interneuron network gamma (PING) or interneuron network gamma (ING) models.^103–105^

The application of MEG as a potential surrogate marker for neurodegenerative states would inform translatable paths to procedural and institutional change to reduce the insidious effects of blast exposure in the military and in contact sports. Biomarkers that can identify individuals at risk of neurodegeneration would be critical in preventing further brain injury, and can play a role in preventative medicine. Along with other safety measures – translated from rodent models that suggest head immobilization during blast exposure can reduce blast-related brain dysfunction^72^ and that jugular compression during breacher training provides preventative utility^36^ – timely integration of longitudinal monitoring with brain-based biomarkers can improve the operational readiness of warfighters and extend the healthy working lifespan of military members and Veterans.

### 4.6 Limitations & future directions

Limitations of this study include the use of a subjective self-reported probe of lifetime blast exposure with the GBEV scale; of course, objective blast exposure data – such as lifetime pressure gauge use – would be advantageous, but due to practical limitations, this data is impossible to capture in our current cohort. Second, multimodal MEG and PET imaging data with Flortaucipir could confirm the presence of tau deposition and spatially colocalized neuronal slowing. Confirmatory tau-slowing associations in a single cohort with suspected CTE would be a step change in our understanding of the relationship between tau-induced neurodegeneration and neural slowing, and confirm the latter as a robust surrogate marker for tauopathy. Third, there were no significant relationships between brain measures and self-reported outcomes – a longitudinal approach, with baseline capture before explosives weapons training, would be crucial in establishing slowing as a marker of neurological functioning, given the between-subject heterogeneity and within-subject stability of brain markers, as well as using objective clinical measures (e.g. neuropsychological testing).^54^

In future work, we plan to examine white matter microstructure in this same cohort, and establish whether white matter damage leads to neural desynchrony, as white matter damage has been reported in contact sport players (football) in the absence of concussions after only a single season of play,^106,107^ as well as MEG tasks in cognitive control, mental flexibility, and memory. Furthermore, there has been major, recent interest in expanding MEG analyses beyond canonical frequency band, periodic analyses as applied here and quantifying the aperiodic component of the neurophysiological signal recorded by MEG.^108^ Studies have shown age-related changes in the aperiodic component,^109^ and aperiodic alterations in AD^110^ and Parkinson’s disease and dementia with Lewy bodies.^111^ Finally, future research should also aim to pair MEG with PET-tau and PET-amyloid as seen in the tauopathy studies mentioned above (e.g., Schoonhoven *et al*.^94^; Schultz *et al*.^95^; García-Colomo *et al*.^96^), as well as other various biomarkers,^35^ including cerebrovascular^112^ and genetic^113^ serum and CSF biomarkers.^114^

### 4.7 Conclusions

Electrophysiological imaging using MEG produced the first evidence of dysregulated neuronal functioning in a cohort with a history of repetitive subconcussions, including military members and Veterans with varying degrees of exposure to repetitive blast overpressure. Notably, these aberrations were observed independently of concussion or traumatic stress history. In addition to regional slowing of activity as indicated by increased delta activity and functional dysconnectivity in key brain hubs and networks, those with worse blast exposure also exhibited worse neurological symptoms, especially in the somatic and cognitive domains, with no blast-related differences in mental health outcomes. MEG has potential utility as a biomarker for neurodegeneration which can translate to procedural and institutional improvements to minimize harm from subconcussions in military blast exposure and in contact sports.

## Data Availability

Defence considerations related to the confidential nature of the human data collected means it is not publicly available.

## Acknowledgements

This research was funded by awards to BTD from the Department of National Defence, and their Innovation for Defence Excellence and Security (IDEaS) program, Defence Research and Development Canada (DRDC), and industry research funding from MYndspan Ltd.

IDEaS is a defence innovation program which invests in research and technology aimed at meeting the demands of the current and future global defence and security environment. Since launching in April 2018, IDEaS has been working with Canadian innovators to develop the S&T landscape, and is helping turn innovative thinking into tangible solutions for the Department of National Defence and the Canadian Armed Forces (DND/CAF), as well as Canadians. Visit the <u>IDEaS web site</u> to get updates on the latest challenges issued by the program.

## References

1. Giza CC, Hovda DA. The new neurometabolic cascade of concussion. Neurosurgery. 2014;75:S24–S33. doi:10.1227/NEU.0000000000000505

2. Bailes JE, Petraglia AL, Omalu BI, Nauman E, Talavage T. Role of subconcussion in repetitive mild traumatic brain injury. J Neurosurg. 2013;119(5):1235–1245. doi:10.3171/2013.7.JNS121822

3. Reynolds BB, Stanton AN, Soldozy S, Goodkin HP, Wintermark M, Druzgal TJ. Investigating the effects of subconcussion on functional connectivity using mass-univariate and multivariate approaches. Brain Imaging Behav. 2018;12(5):1332–1345. doi:10.1007/s11682-017-9790-z

4. Dashnaw ML, Petraglia AL, Bailes JE. An overview of the basic science of concussion and subconcussion: Where we are and where we are going. Neurosurg Focus. 2012;33(6):1–9. doi:10.3171/2012.10.FOCUS12284

5. Stein TD, Alvarez VE, McKee AC. Concussion in Chronic Traumatic Encephalopathy. Curr Pain Headache Rep. 2015;19(10):47. doi:10.1007/s11916-015-0522-z

6. Daneshvar DH, Nair ES, Baucom ZH, et al. Leveraging football accelerometer data to quantify associations between repetitive head impacts and chronic traumatic encephalopathy in males. Nat Commun. 2023;14(1):3470. doi:10.1038/s41467-023-39183-0

7. Vartanian O, Rhind SG, Nakashima A, et al. Blast effects on post-concussive and mental health outcomes: Data from Canadian Armed Forces breachers and snipers. J Mil Veteran Fam Heal. 2022;8(s2):82–96. doi:10.3138/jmvfh-2021-0122

8. Vartanian O, Tenn C, Rhind SG, et al. Blast in context: The neuropsychological and neurocognitive effects of long-term occupational exposure to repeated low-level explosives on Canadian Armed Forces’ breaching instructors and range staff. Front Neurol. 2020;11:588531. doi:10.3389/fneur.2020.588531

9. Nakashima A, Vartanian O, Rhind SG, King K, Tenn C, Jetly CR. Repeated occupational exposure to low-level blast in the Canadian Armed Forces: Effects on hearing, balance, and ataxia. Mil Med. 2022;187(1/2):e201–e208. doi:10.1093/milmed/usaa439

10. Vartanian O, Coady L, Blackler K, Fraser B, Cheung B. Neuropsychological, neurocognitive, vestibular, and neuroimaging correlates of exposure to repetitive low-level blast waves: Evidence from four nonoverlapping samples of Canadian breachers. Mil Med. 2021;186(3/4):e393–e400. doi:10.1093/milmed/usaa332

11. Robinson ME, McKee AC, Salat DH, et al. Positron emission tomography of tau in Iraq and Afghanistan Veterans with blast neurotrauma. NeuroImage Clin. 2019;21:101651. doi:10.1016/j.nicl.2019.101651

12. Kong LZ, Zhang RL, Hu SH, Lai JB. Military traumatic brain injury: a challenge straddling neurology and psychiatry. Mil Med Res. 2022;9(1):2. doi:10.1186/s40779-021-00363-y

13. Babcock KJ, Abdolmohammadi B, Kiernan PT, et al. Interface astrogliosis in contact sport head impacts and military blast exposure. Acta Neuropathol Commun. 2022;10:52. doi:10.1186/s40478-022-01358-z

14. Priemer DS, Iacono D, Rhodes CH, Olsen CH, Perl DP. Chronic Traumatic Encephalopathy in the brains of military personnel. N Engl J Med. 2022;386(23):2169–2177. doi:10.1056/nejmoa2203199

15. Huang CX, Li YH, Lu W, et al. Positron emission tomography imaging for the assessment of mild traumatic brain injury and chronic traumatic encephalopathy: recent advances in radiotracers. Neural Regen Res. 2022;17(1):74–81. doi:10.4103/1673-5374.314285

16. Varlow C, Vasdev N. Evaluation of tau radiotracers in Chronic Traumatic Encephalopathy. J Nucl Med. 2023;64(3):460–465. doi:10.2967/jnumed.122.264404

17. Coomans EM, Schoonhoven DN, Tuncel H, et al. In vivo tau pathology is associated with synaptic loss and altered synaptic function. Alzheimer’s Res Ther. 2021;13(1):35. doi:10.1186/s13195-021-00772-0

18. Franzmeier N, Brendel M, Beyer L, et al. Tau deposition patterns are associated with functional connectivity in primary tauopathies. Nat Commun. 2022;13(1):1362. doi:10.1038/s41467-022-28896-3

19. Uhlhaas PJ, Singer W. Neural synchrony in brain disorders: Relevance for cognitive dysfunctions and pathophysiology. Neuron. 2006;52(1):155–168. doi:10.1016/j.neuron.2006.09.020

20. Dunkley BT. Differential intrinsic coupling modes in psychological and physical trauma. Front Psychiatry. 2015;6:140. doi:10.3389/fpsyt.2015.00140

21. Zhang J, Emami Z, Safar K, et al. Teasing apart trauma: neural oscillations differentiate individual cases of mild traumatic brain injury from post-traumatic stress disorder even when symptoms overlap. Transl Psychiatry. 2021;11(1):345. doi:10.1038/s41398-021-01467-8

22. Rier L, Zamyadi R, Zhang J, et al. Mild traumatic brain injury impairs the coordination of intrinsic and motor-related neural dynamics. NeuroImage Clin. 2021;32:102841. doi:10.1016/j.nicl.2021.102841

23. Allen CM, Halsey L, Topcu G, et al. Magnetoencephalography abnormalities in adult mild traumatic brain injury: A systematic review. NeuroImage Clin. 2021;31:102697. doi:10.1016/j.nicl.2021.102697

24. Huang MX, Nichols S, Baker DG, et al. Single-subject-based whole-brain MEG slow-wave imaging approach for detecting abnormality in patients with mild traumatic brain injury. NeuroImage Clin. 2014;5:109–119. doi:10.1016/j.nicl.2014.06.004

25. Kaltiainen H, Liljeström M, Helle L, et al. Mild traumatic brain injury affects cognitive processing and modifies oscillatory brain activity during attentional tasks. J Neurotrauma. 2019;36(14):2222–2232. doi:10.1089/neu.2018.6306

26. Dunkley BT, Da Costa L, Bethune A, et al. Low-frequency connectivity is associated with mild traumatic brain injury. NeuroImage Clin. 2015;7:611–621. doi:10.1016/j.nicl.2015.02.020

27. Wiesman AI, da Silva Castanheira J, Degroot C, Fon EA, Baillet S, Network QP. Adverse and compensatory neurophysiological slowing in Parkinson’s disease. Prog Neurobiol. 2023;231:102538. doi:10.1016/j.pneurobio.2023.102538

28. Wiesman AI, Murman DL, Losh RA, et al. Spatially resolved neural slowing predicts impairment and amyloid burden in Alzheimer’s disease. Brain. 2022;145(6):2177–2189. doi:10.1093/BRAIN/AWAB430

29. Modica LCM, Egnoto MJ, Statz JK, Carr W, Ahlers ST. Development of a blast exposure estimator from a department of defense-wide survey study on military service members. J Neurotrauma. 2021;38(12):1654–1661. doi:10.1089/neu.2020.7405

30. Lange RT, French LM, Lippa SM, et al. High lifetime blast exposure using the blast exposure threshold survey is associated with worse warfighter brain health following mild traumatic brain injury. J Neurotrauma. 2024;41(1/2):186–198. doi:10.1089/neu.2023.0133

31. Mu W, Catenaccio E, Lipton ML. Neuroimaging in blast-related mild traumatic brain injury. J Head Trauma Rehabil. 2017;32(1):55–69. doi:10.1097/HTR.0000000000000213

32. Huang MX, Harrington DL, Robb Swan A, et al. Resting-state magnetoencephalography reveals different patterns of aberrant functional connectivity in combat-related mild traumatic brain injury. J Neurotrauma. 2017;34(7):1412–1426. doi:10.1089/neu.2016.4581

33. Huang M, Risling M, Baker DG. The role of biomarkers and MEG-based imaging markers in the diagnosis of post-traumatic stress disorder and blast-induced mild traumatic brain injury. Psychoneuroendocrinology. 2016;63:398–409. doi:10.1016/j.psyneuen.2015.02.008

34. Rowland JA, Stapleton-Kotloski JR, Martindale SL, et al. Alterations in the topology of functional connectomes are associated with Post-Traumatic Stress Disorder and blast-related mild traumatic brain injury in combat Veterans. J Neurotrauma. 2021;38(22):3086–3096. doi:10.1089/neu.2020.7450

35. Wilde EA, Wanner IB, Kenney K, et al. A framework to advance biomarker development in the diagnosis, outcome prediction, and treatment of traumatic brain injury. J Neurotrauma. 2022;39(7-8):436–457. doi:10.1089/neu.2021.0099

36. Yuan W, Barber Foss KD, Dudley J, et al. Impact of low-level blast eposure on brain function after a one-day tactile training and the ameliorating effect of a jugular vein compression neck collar device. J Neurotrauma. 2019;36(5):721–734. doi:10.1089/neu.2018.5737

37. Abbas K, Shenk TE, Poole VN, et al. Alteration of default mode network in high school football athletes due to repetitive subconcussive mild traumatic brain injury: A resting-state functional magnetic resonance imaging study. Brain Connect. 2015;5(2):91–101. doi:10.1089/brain.2014.0279

38. Stone JR, Avants BB, Tustison NJ, et al. Neurological effects of repeated blast exposure in Special Operations personnel. J Neurotrauma. Published online 2024. doi:10.1089/neu.2023.0309

39. Akin FW, Murnane OD, Hall CD, Riska KM. Vestibular consequences of mild traumatic brain injury and blast exposure: a review. Brain Inj. 2017;31(9):1188–1194. doi:10.1080/02699052.2017.1288928

40. Franke LM, Walker WC, Cifu DX, Ochs AL, Lew HL. Sensorintegrative dysfunction underlying vestibular disorders after traumatic brain injury: A review. J Rehabil Res Dev. 2012;49(7):985–994. doi:10.1682/JRRD.2011.12.0250

41. Elsayed NM. Toxicology of blast overpressure. Toxicology. 1997;121(1):1–15. doi:10.1016/S0300-483X(97)03651-2

42. Mayorga MA. The pathology of primary blast overpressure injury. Toxicology. 1997;121(1):17–28. doi:10.1016/S0300-483X(97)03652-4

43. Edlow BL, Bodien YG, Baxter T, et al. Long-term effects of repeated blast exposure in United States Special Operations Forces personnel: A pilot study protocol. J Neurotrauma. 2022;39(19-20):1391–1407. doi:10.1089/neu.2022.0030

44. Spitzer RL, Kroenke K, Williams JW, Löwe B. A brief measure for assessing Generalized Anxiety Disorder: The GAD-7. Arch Intern Med. 2006;166(10):1092–1097. doi:10.1001/archinte.166.10.1092

45. Kroenke K, Spitzer RL, Williams JBW. The PHQ-9: Validity of a brief depression severity measure. J Gen Intern Med. 2001;16(9):606–613. doi:10.1046/j.1525-1497.2001.016009606.x

46. Weathers FW, Litz BT, Herman DS, Huska JA, Keane TM. PTSD Checklist--Military Version. APA PsycTests; 1993. doi:10.1037/t05198-000

47. Echemendia RJ, Meeuwisse W, Mccrory P, et al. The Sport Concussion Assessment Tool 5th Edition (SCAT5): Background and rationale. Br J Sport Med. 2017;51:848–850. doi:10.1136/bjsports-2017-097506

48. Gioia GA, Collins M, Isquith PK. Improving identification and diagnosis of mild traumatic brain injury with evidence: psychometric support for the acute concussion evaluation. J Head Trauma Rehabil. 2008;23(4):230–242. doi:10.1097/01.HTR.0000327255.38881.ca

49. Schnurr PP, Vielhauer MJ, Weathers F, Findler MN. Brief Trauma Questionnaire (BTQ). APA PsycTests; 2009. doi:10.1037/t07488-000

50. Oostenveld R, Fries P, Maris E, Schoffelen JM. FieldTrip: Open source software for advanced analysis of MEG, EEG, and invasive electrophysiological data. Comput Intell Neurosci. 2011;2011:156869. doi:10.1155/2011/156869

51. Tzourio-Mazoyer N, Landeau B, Papathanassiou D, et al. Automated anatomical labeling of activations in SPM using a macroscopic anatomical parcellation of the MNI MRI single-subject brain. Neuroimage. 2002;15(1):273–289. doi:10.1006/nimg.2001.0978

52. Van Veen BD, van Drongelen W, Yuchtman M, Suzuki A. Localization of brain electrical activity via linearly constrained minimum variance spatial filtering. IEEE Trans Biomed Eng. 1997;44(9):867–880. doi:10.1109/10.623056

53. Colclough GL, Brookes MJ, Smith SM, Woolrich MW. A symmetric multivariate leakage correction for MEG connectomes. Neuroimage. 2015;117:439–448. doi:10.1016/j.neuroimage.2015.03.071

54. Lew BJ, Fitzgerald EE, Ott LR, Penhale SH, Wilson TW. Three-year reliability of MEG resting-state oscillatory power. Neuroimage. 2021;243:118516. doi:10.1016/j.neuroimage.2021.118516

55. Brookes MJ, Woolrich MW, Barnes GR. Measuring functional connectivity in MEG: A multivariate approach insensitive to linear source leakage. Neuroimage. 2012;63(2):910–920. doi:10.1016/j.neuroimage.2012.03.048

56. Dunkley BT, Urban K, Da Costa L, Wong SM, Pang EW, Taylor MJ. Default mode network oscillatory coupling is increased following concussion. Front Neurol. 2018;9:280. doi:10.3389/fneur.2018.00280

57. Safar K, Zhang J, Emami Z, Gharehgazlou A, Ibrahim G, Dunkley BT. Mild traumatic brain injury is associated with dysregulated neural network functioning in children and adolescents. Brain Commun. 2021;3(2):fcab044. doi:10.1093/braincomms/fcab044

58. Colclough GL, Woolrich MW, Tewarie PK, Brookes MJ, Quinn AJ, Smith SM. How reliable are MEG resting-state connectivity metrics? Neuroimage. 2016;138:284–293. doi:10.1016/j.neuroimage.2016.05.070

59. Chao-Gan Y, Yu-Feng Z. DPARSF: A MATLAB toolbox for “pipeline” data analysis of resting-state fMRI. Front Syst Neurosci. 2010;4:13. doi:10.3389/fnsys.2010.00013

60. Penny W, Friston K, Ashburner J, Kiebel S, Nichols T. Statistical Parametric Mapping: The Analysis of Functional Brain Images. Elsevier/Academic Press; 2007. doi:10.1016/B978-0-12-372560-8.X5000-1

61. Friston KJ, Frith CD, Frackowiak RSJ, Turner R. Characterizing dynamic brain responses with fMRI: A multivariate approach. Neuroimage. 1995;2(2):166–172. doi:10.1006/nimg.1995.1019

62. Greicius MD, Krasnow B, Reiss AL, Menon V. Functional connectivity in the resting brain: A network analysis of the default mode hypothesis. Proc Natl Acad Sci U S A. 2003;100(1):253–258. doi:10.1073/pnas.0135058100

63. Fox MD, Snyder AZ, Vincent JL, Corbetta M, Van Essen DC, Raichle ME. The human brain is intrinsically organized into dynamic, anticorrelated functional networks. Proc Natl Acad Sci U S A. 2005;102(27):9673–9678. doi:10.1073/pnas.0504136102

64. Fransson P. Spontaneous low-frequency BOLD signal fluctuations: An fMRI investigation of the resting-state default mode of brain function hypothesis. Hum Brain Mapp. 2005;26(1):15–29. doi:10.1002/hbm.20113

65. Clare Kelly AM, Uddin LQ, Biswal BB, Castellanos FX, Milham MP. Competition between functional brain networks mediates behavioral variability. Neuroimage. 2008;39(1):527–537. doi:10.1016/j.neuroimage.2007.08.008

66. Lu H, Zuo Y, Gu H, et al. Synchronized delta oscillations correlate with the resting-state functional MRI signal. Proc Natl Acad Sci U S A. 2007;104(46):18265–18269. doi:10.1073/pnas.0705791104

67. Biswal B, Yetkin FZ, Haughton VM, Hyde JS. Functional connectivity in the motor cortex of resting human brain using echo-planar MRI. Magn Reson Med. 1995;34(4):537–541. doi:10.1002/mrm.1910340409

68. Lowe MJ, Mock BJ, Sorenson JA. Functional connectivity in single and multislice echoplanar imaging using resting-state fluctuations. Neuroimage. 1998;7(2):119–132. doi:10.1006/nimg.1997.0315

69. Tewarie P, Hillebrand A, van Dellen E, et al. Structural degree predicts functional network connectivity: A multimodal resting-state fMRI and MEG study. Neuroimage. 2014;97:296–307. doi:10.1016/j.neuroimage.2014.04.038

70. Zalesky A, Fornito A, Bullmore ET. Network-based statistic: Identifying differences in brain networks. Neuroimage. 2010;53(4):1197–1207. doi:10.1016/j.neuroimage.2010.06.041

71. Bugay V, Bozdemir E, Vigil FA, et al. A mouse model of repetitive blast traumatic brain injury reveals post-trauma seizures and increased neuronal excitability. J Neurotrauma. 2020;37(2):248–261. doi:10.1089/neu.2018.6333

72. Goldstein LE, Fisher AM, Tagge CA, et al. Chronic traumatic encephalopathy in blast-exposed military veterans and a blast neurotrauma mouse model. Sci Transl Med. 2012;4(134):134ra60. doi:10.1126/scitranslmed.3003716

73. Aravind A, Ravula AR, Chandra N, Pfister BJ. Behavioral deficits in animal models of blast traumatic brain injury. Front Neurol. 2020;11:990. doi:10.3389/fneur.2020.00990

74. Siedhoff HR, Chen S, Song H, et al. Perspectives on primary blast injury of the brain: Translational insights into non-inertial low-intensity blast injury. Front Neurol. 2022;12:818169. doi:10.3389/fneur.2021.818169

75. Dickerson MR, Murphy SF, Urban MJ, White Z, VandeVord PJ. Chronic anxiety- and depression-like behaviors are associated with glial-driven pathology following repeated blast induced neurotrauma. Front Behav Neurosci. 2021;15:787475. doi:10.3389/fnbeh.2021.787475

76. Perez Garcia G, De Gasperi R, Gama Sosa MA, et al. Laterality and region-specific tau phosphorylation correlate with PTSD-related behavioral traits in rats exposed to repetitive low-level blast. Acta Neuropathol Commun. 2021;9(1):33. doi:10.1186/s40478-021-01128-3

77. Hernandez A, Tan C, Plattner F, et al. Exposure to mild blast forces induces neuropathological effects, neurophysiological deficits and biochemical changes. Mol Brain. 2018;11(1):64. doi:10.1186/s13041-018-0408-1

78. Sajja VSSS, Hubbard WB, Hall CS, Ghoddoussi F, Galloway MP, VandeVord PJ. Enduring deficits in memory and neuronal pathology after blast-induced traumatic brain injury. Sci Rep. 2015;5:15075. doi:10.1038/srep15075

79. Mckee AC, Daneshvar DH. The neuropathology of traumatic brain injury. Handb Clin Neurol. 2015;127:45–66. doi:10.1016/B978-0-444-52892-6.00004-0

80. Jellinger KA. Basic mechanisms of neurodegeneration: A critical update. J Cell Mol Med. 2010;14(3):457–487. doi:10.1111/j.1582-4934.2010.01010.x

81. Guzman-Martinez L, Maccioni RB, Andrade V, Navarrete LP, Pastor MG, Ramos-Escobar N. Neuroinflammation as a common feature of neurodegenerative disorders. Front Pharmacol. 2019;10:1008. doi:10.3389/fphar.2019.01008

82. Huang MX, Theilmann RJ, Robb A, et al. Integrated imaging approach with MEG and DTI to detect mild traumatic brain injury in military and civilian patients. J Neurotrauma. 2009;26(8):1213–1226. doi:10.1089/neu.2008.0672

83. Giustiniani A, Danesin L, Bozzetto B, Macina A, Benavides-Varela S, Burgio F. Functional changes in brain oscillations in dementia: a review. Rev Neurosci. 2023;34(1):25–47. doi:10.1515/revneuro-2022-0010

84. Leech R, Sharp DJ. The role of the posterior cingulate cortex in cognition and disease. Brain. 2014;137(1):12–32. doi:10.1093/brain/awt162

85. van den Heuvel MP, Sporns O. Rich-club organization of the human connectome. J Neurosci. 2011;31(44):15775–15786. doi:10.1523/JNEUROSCI.3539-11.2011

86. Baggio HC, Segura B, Junque C, de Reus MA, Sala-Llonch R, Van den Heuvel MP. Rich club organization and cognitive performance in healthy older participants. J Cogn Neurosci. 2015;27(9):1801–1810. doi:10.1162/jocn

87. Zhou Y, Milham MP, Lui YW, et al. Default-mode network disruption in mild traumatic brain injury. Radiology. 2012;265(3):882–892. doi:10.1148/radiol.12120748

88. Zhang H, Dai R, Qin P, et al. Posterior cingulate cross-hemispheric functional connectivity predicts the level of consciousness in traumatic brain injury. Sci Rep. 2017;7(1):387. doi:10.1038/s41598-017-00392-5

89. DeSimone JC, Davenport EM, Urban J, et al. Mapping default mode connectivity alterations following a single season of subconcussive impact exposure in youth football. Hum Brain Mapp. 2021;42(8):2529–2545. doi:10.1002/hbm.25384

90. Buzsáki G, Schomburg EW. What does gamma coherence tell us about inter-regional neural communication? Nat Neurosci. 2015;18(4):484–489. doi:10.1038/nn.3952

91. Wu JW, Hussaini SA, Bastille IM, et al. Neuronal activity enhances tau propagation and tau pathology in vivo. Nat Neurosci. 2016;19(8):1085–1095. doi:10.1038/nn.4328

92. Pooler AM, Phillips EC, Lau DHW, Noble W, Hanger DP. Physiological release of endogenous tau is stimulated by neuronal activity. EMBO Rep. 2013;14(4):389–394. doi:10.1038/embor.2013.15

93. Yamada K, Holth JK, Liao F, et al. Neuronal activity regulates extracellular tau in vivo. J Exp Med. 2014;211(3):387–393. doi:10.1084/jem.20131685

94. Schoonhoven DN, Coomans EM, Millán AP, et al. Tau protein spreads through functionally connected neurons in Alzheimer’s disease: a combined MEG/PET study. Brain. 2023;146(10):4040–4054. doi:10.1093/brain/awad189

95. Schultz AP, Chhatwal JP, Hedden T, et al. Phases of hyperconnectivity and hypoconnectivity in the default mode and salience networks track with amyloid and tau in clinically normal individuals. J Neurosci. 2017;37(16):4323–4331. doi:10.1523/JNEUROSCI.3263-16.2017

96. García-Colomo A, Nebreda A, Carrasco-Gómez M, et al. Longitudinal changes in the functional connectivity of individuals at risk of Alzheimer’s disease. GeroSciencecience. Published online 2024. doi:10.1007/s11357-023-01036-5

97. Stuss DT, Knight RT. Principles of Frontal Lobe Function. Online edi. Oxford Academic; 2013.

98. Burgess N, Maguire EA, O’Keefe J. The human hippocampus and spatial and episodic memory. Neuron. 2002;35(4):625–641. doi:10.1016/S0896-6273(02)00830-9

99. Postle BR. The hippocampus, memory, and consciousness. In: The Neurology of Consciousness: Cognitive Neuroscience and Neuropathology. Second edi. Academic Press; 2015:349–363. doi:10.1016/B978-0-12-800948-2.00021-2

100. Baddeley A, Jarrold C, Vargha-Khadem F. Working memory and the hippocampus. J Cogn Neurosci. 2011;23(12):3855-3861. doi:10.1162/jocn_a_00066

101. Lisman J, Buzsáki G, Eichenbaum H, Nadel L, Rangananth C, Redish AD. Viewpoints: how the hippocampus contributes to memory, navigation and cognition. Nat Neurosci. 2017;20(11):1434–1447. doi:10.1038/nn.4661

102. Rubin RD, Watson PD, Duff MC, Cohen NJ. The role of the hippocampus in flexible cognition and social behavior. Front Hum Neurosci. 2014;8(SEP):1–15. doi:10.3389/fnhum.2014.00742

103. Buzsáki G, Wang XJ. Mechanisms of gamma oscillations. Annu Rev Neurosci. 2012;35:203–225. doi:10.1146/annurev-neuro-062111-150444.Mechanisms

104. Börgers C, Epstein S, Kopell NJ. Background gamma rhythmicity and attention in cortical local circuits: a computational study. Proc Natl Acad Sci U S A. 2005;102(19):7002–7007. doi:10.1073/pnas.0502366102

105. Guan A, Wang S, Huang A, et al. The role of gamma oscillations in central nervous system diseases: Mechanism and treatment. Front Cell Neurosci. 2022;16:962957. doi:10.3389/fncel.2022.962957

106. Davenport EM, Whitlow CT, Urban JE, et al. Abnormal white matter integrity related to head impact exposure in a season of high school varsity football. J Neurotrauma. 2014;31(19):1617–1624. doi:10.1089/neu.2013.3233

107. Bahrami N, Sharma D, Rosenthal S, et al. Subconcussive head impact exposure and white matter tract changes over a single season of youth football. Radiology. 2016;281(3):919–926. doi:10.1148/radiol.2016160564

108. Donoghue T, Haller M, Peterson EJ, et al. Parameterizing neural power spectra into periodic and aperiodic components. Nat Neurosci. 2020;23(12):1655–1665. doi:10.1038/s41593-020-00744-x

109. Hill AT, Clark GM, Bigelow FJ, Lum JAG, Enticott PG. Periodic and aperiodic neural activity displays age-dependent changes across early-to-middle childhood. Dev Cogn Neurosci. 2022;54:101076. doi:10.1016/j.dcn.2022.101076

110. van Nifterick AM, Mulder D, Duineveld DJ, et al. Resting-state oscillations reveal disturbed excitation–inhibition ratio in Alzheimer’s disease patients. Sci Rep. 2023;13(1):7419. doi:10.1038/s41598-023-33973-8

111. Rosenblum Y, Shiner T, Bregman N, et al. Decreased aperiodic neural activity in Parkinson’s disease and dementia with Lewy bodies. J Neurol. 2023;270(8):3958–3969. doi:10.1007/s00415-023-11728-9

112. Baker TL, Agoston D V., Brady RD, et al. Targeting the cerebrovascular system: Next-generation biomarkers and treatment for mild traumatic brain injury. Neuroscientist. 2022;28(6):594–612. doi:10.1177/10738584211012264

113. Taskina D, Zhu C, Schwab N, Hazrati LN. Brain pathology and symptoms linked to concussion history: beyond chronic traumatic encephalopathy. Brain Commun. Published online February 26, 2024:fcad314. doi:10.1093/braincomms/fcad314

114. Li G, Iliff J, Shofer J, et al. CSF β-amyloid and tau biomarker changes in Veterans with mild traumatic brain injury. Neurology. 2024;102(7):e209197. doi:10.1212/WNL.0000000000209197

