## Supplementary material for "Repetitive subconcussion results in disrupted neural activity independent of concussion history"

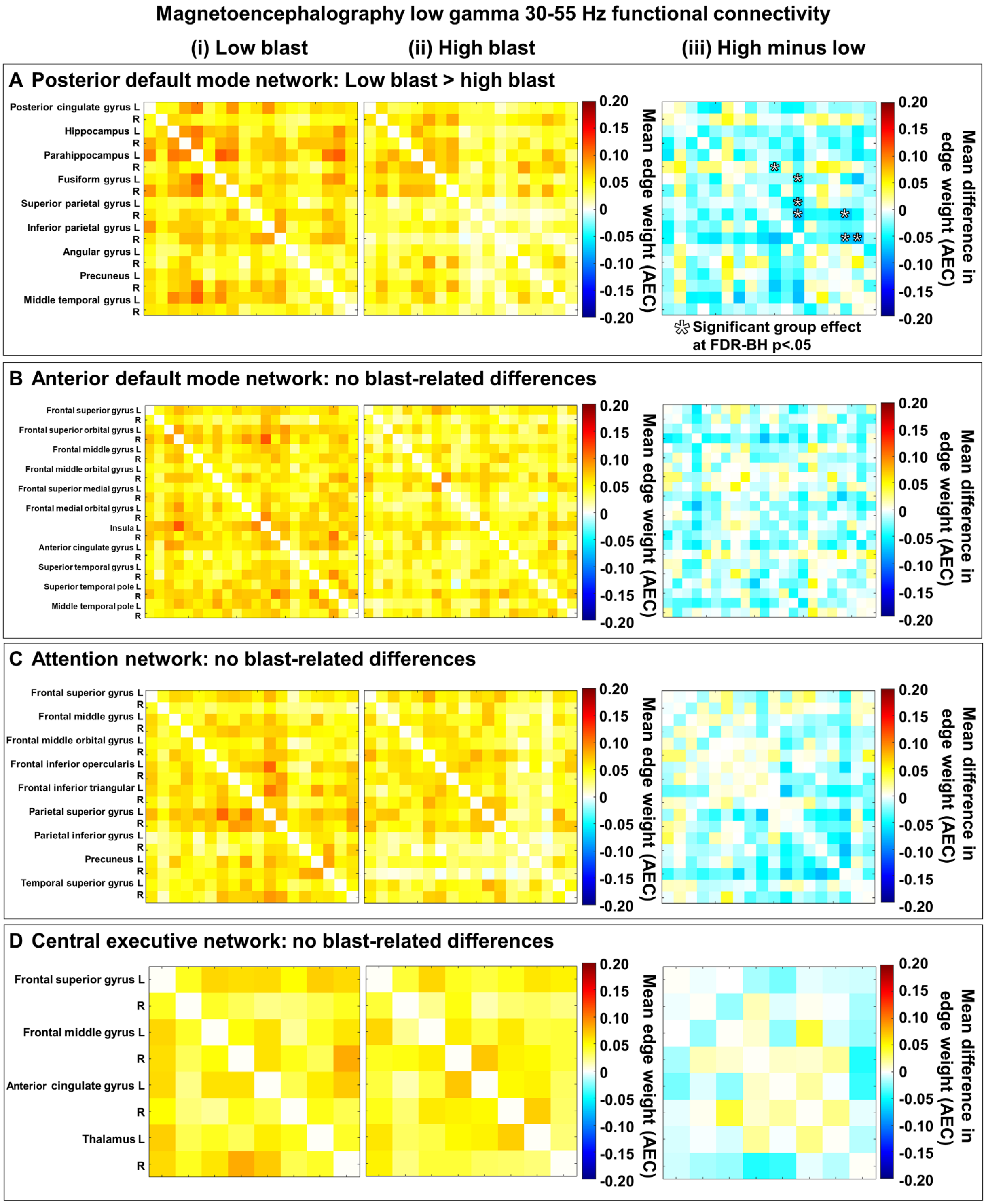

**Figure S1:** Functional connectivity matrices scaled by mean edge weight (AEC) measured by resting-state magnetoencephalography (MEG) at low gamma (30-55 Hz) range in the (**A**) posterior (pDMN) and (**B**) anterior default mode networks (aDMN), (**C**) attention network (AN), and (**D**) central executive control network (CEN) with the mean connectivity for individuals with (**i**) low blast exposure and (**ii**) high blast exposure, and (**iii**) the group difference. (**Aiii**) In the pDMN, there was significantly lower connectivity in the high blast (**Aii**) relative to the low blast exposure group (**Ai**) for 7 nodes and 7 edges at low gamma; the node with the highest degree was the right inferior parietal. There were no blast-related differences in (**B**) aDMN, (**C**) AN, or (**D**) CEN functional connectivity as measured by MEG at low gamma. Warm elements in the (**i, ii**) group mean matrices indicate higher connectivity and cool colours indicate lower connectivity; warm elements in the (**iii**) group difference matrices indicate higher connectivity in the high blast group and cool colours indicate lower connectivity in the high blast group.

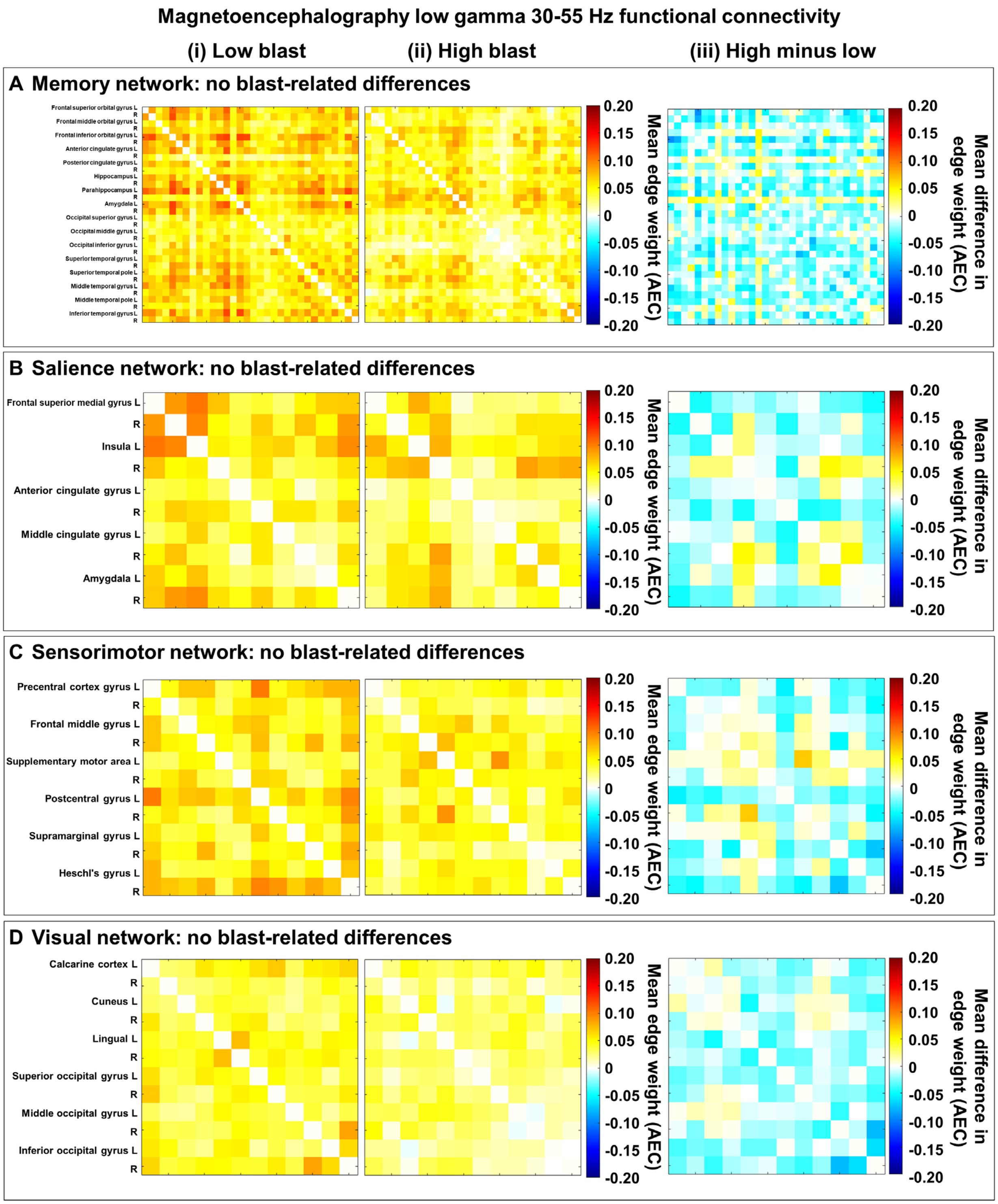


**Figure S2:** Functional connectivity matrices scaled by mean edge weight (AEC) measured by resting-state magnetoencephalography (MEG) at low gamma (30-55 Hz) range in the (**A**) memory network (MN), (**B**) salience network (SN), (**C**) sensorimotor network (SMN), and (**D**) visual network (VN) with the mean connectivity for individuals with (**i**) low blast exposure and (**ii**) high blast exposure, and (**iii**) the group difference.

There were no blast-related differences in (**A**) MN, (**B**) SN, (**C**) SMN, or (**D**) VN functional connectivity as measured by MEG at low gamma. Warm elements in the (**i, ii**) group mean matrices indicate higher connectivity and cool colours indicate lower connectivity; warm elements in the (**iii**) group difference matrices indicate higher connectivity in the high blast group and cool colours indicate lower connectivity in the high blast group.

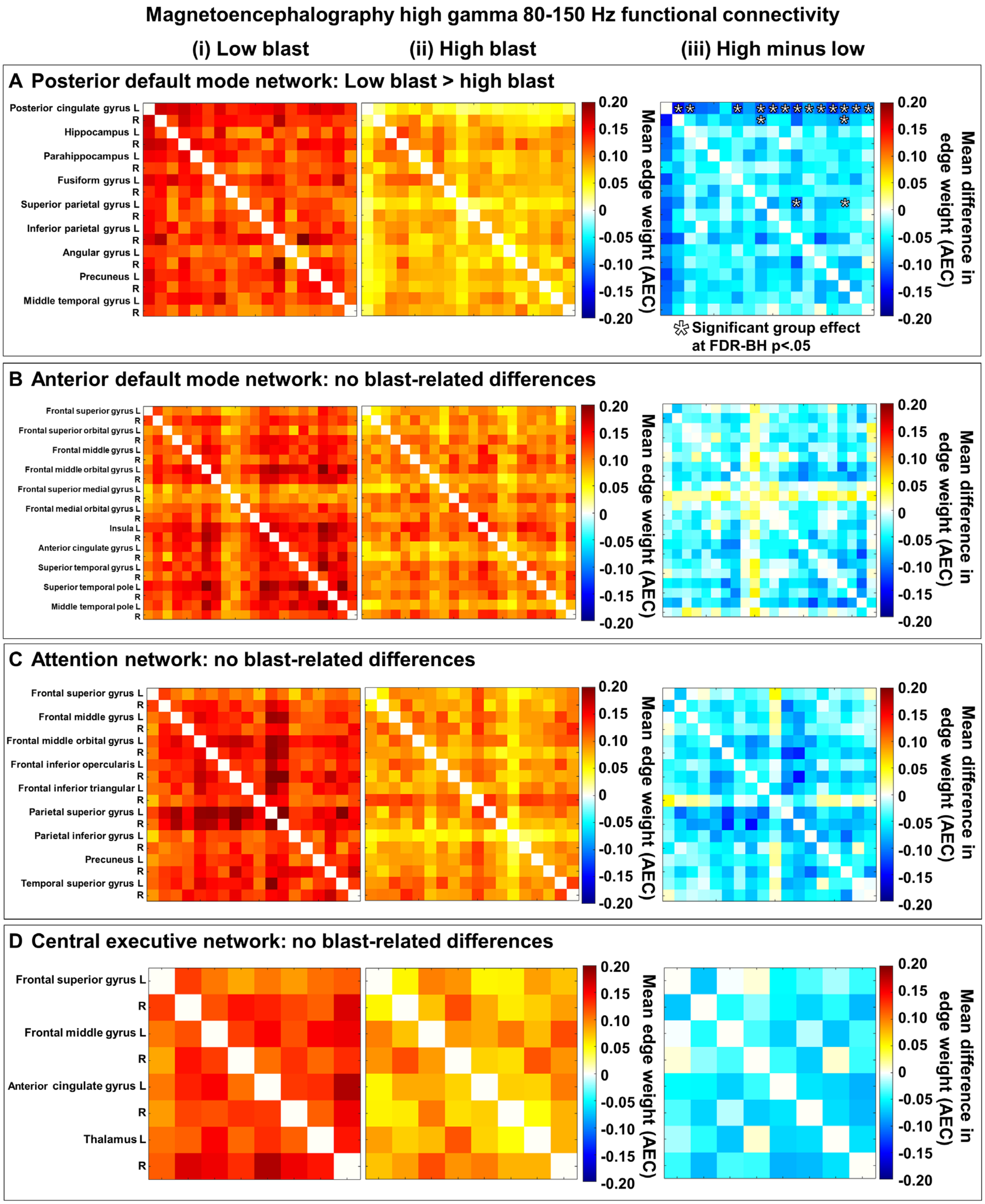


**Figure S3:** Functional connectivity matrices scaled by mean edge weight (AEC) measured by resting-state magnetoencephalography (MEG) at high gamma (80-150 Hz) range in the (**A**) posterior (pDMN) and (**B**) anterior default mode networks (aDMN), (**C**) attention network (AN), and (**D**) central executive control network (CEN) with the mean connectivity for individuals with (**i**) low blast exposure and (**ii**) high blast exposure, and (**iii**) the group difference. (**Aiii**) In the pDMN, there was significantly lower connectivity in the high blast (**Aii**) relative to the low blast exposure group (**Ai**) for 14 nodes and 17 edges at high gamma; the node with the highest degree was the left posterior cingulate. There were no blast-related differences in (**B**) aDMN, (**C**) AN, or (**D**) CEN functional connectivity as measured by MEG at high gamma. Warm elements in the (**i, ii**) group mean matrices indicate higher connectivity and cool colours indicate lower connectivity; warm elements in the (**iii**) group difference matrices indicate higher connectivity in the high blast group and cool colours indicate lower connectivity in the high blast group.

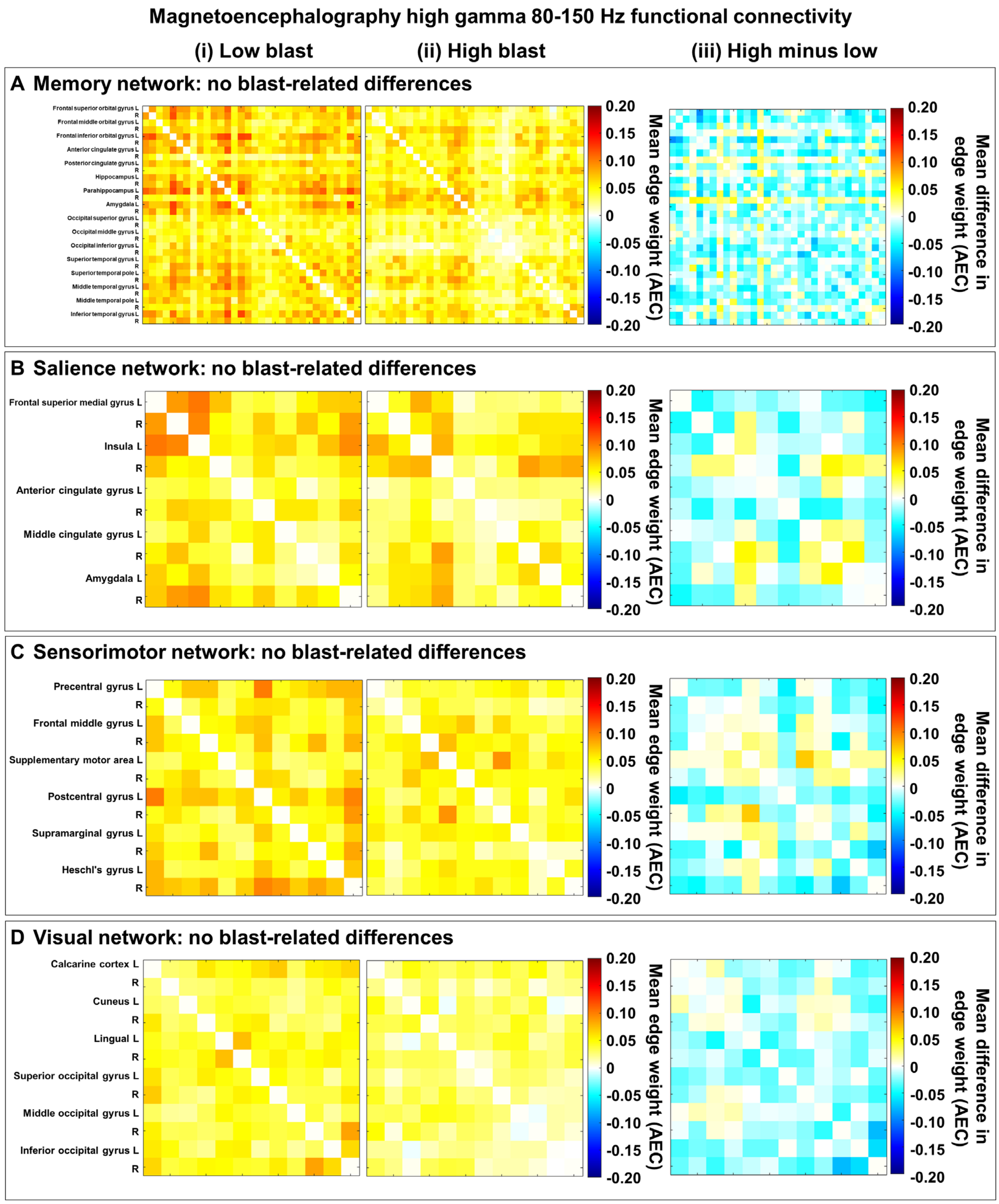


**Figure S4:** Functional connectivity matrices scaled by mean edge weight (AEC) measured by resting-state magnetoencephalography (MEG) at high gamma (80-150 Hz) range in the (**A**) memory network (MN), (**B**) salience network (SN), (**C**) sensorimotor network (SMN), and (**D**) visual network (VN) with the mean connectivity for individuals with (**i**) low blast exposure and (**ii**) high blast exposure, and (**iii**) the group difference.

There were no blast-related differences in (**A**) MN, (**B**) SN, (**C**) SMN, or (**D**) VN functional connectivity as measured by MEG at high gamma. Warm elements in the (**i, ii**) group mean matrices indicate higher connectivity and cool colours indicate lower connectivity; warm elements in the (**iii**) group difference matrices indicate higher connectivity in the high blast group and cool colours indicate lower connectivity in the high blast group.

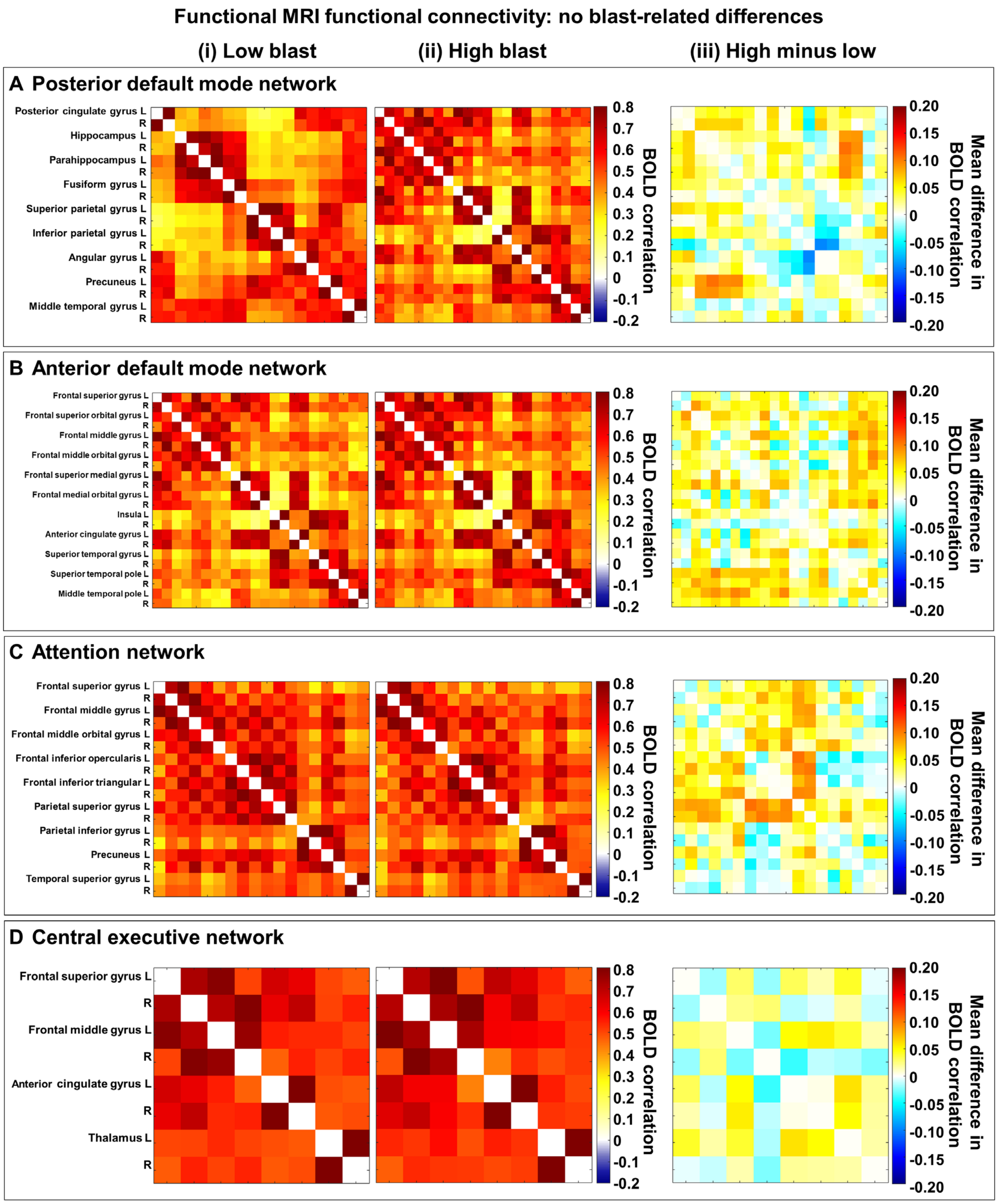


**Figure S5:** Functional connectivity matrices scaled by BOLD correlations indicated by resting-state functional MRI in the (**A**) posterior (pDMN) and (**B**) anterior default mode networks (aDMN), (**C**) attention network (AN), and (**D**) central executive control network (CEN) with the mean connectivity for individuals with (**i**) low blast exposure and (**ii**) high blast exposure, and (**iii**) the group difference. There were no blast-related differences in

(**A**) pDMN, (**B**) aDMN, (**C**) AN, or (**D**) CEN functional connectivity as measured by functional MRI. Warm elements in the (**i, ii**) group mean matrices indicate higher connectivity and cool colours indicate lower connectivity; warm elements in the (**iii**) group difference matrices indicate higher connectivity in the high blast group and cool colours indicate lower connectivity in the high blast group.

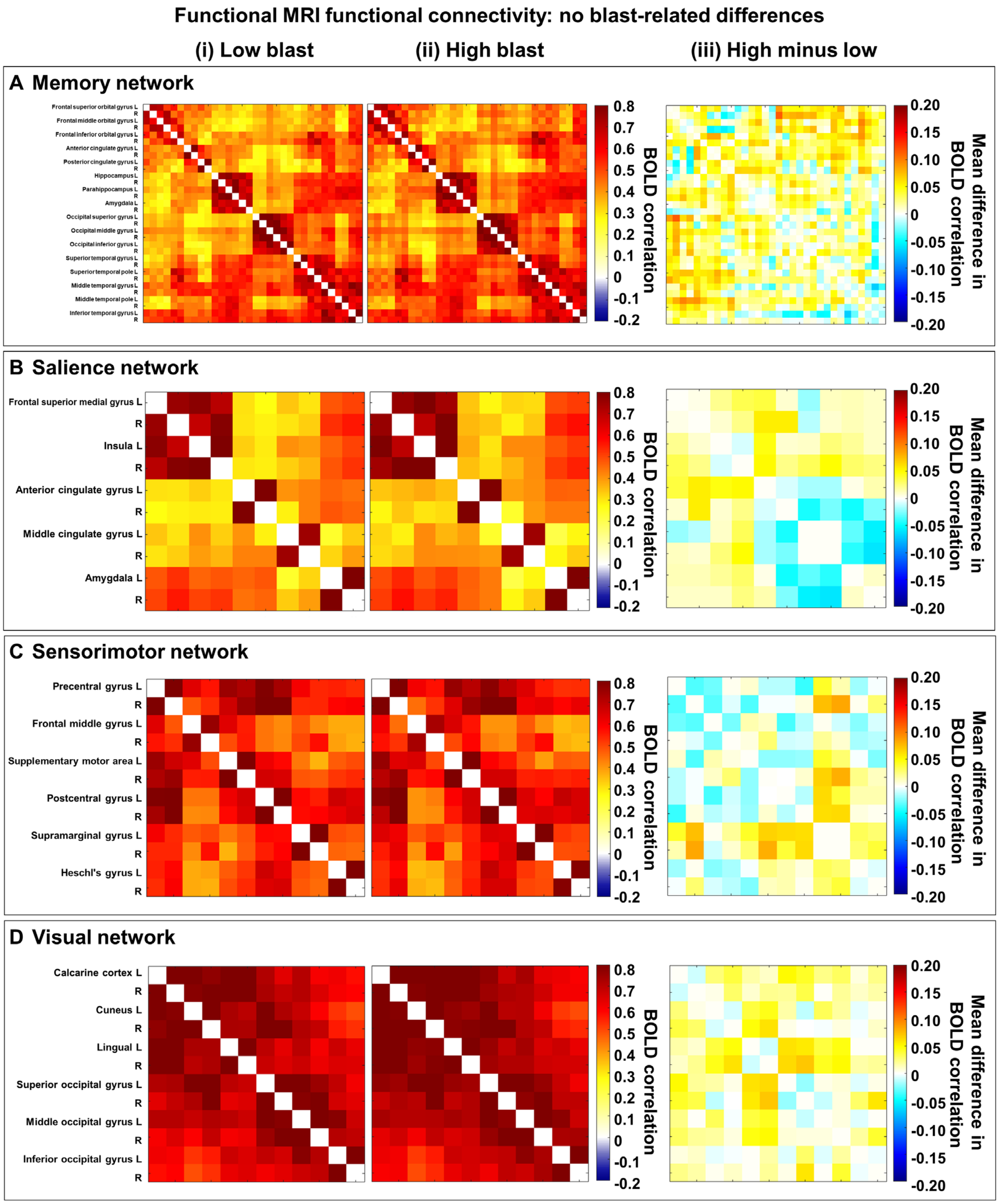


**Figure S6:** Functional connectivity matrices scaled by BOLD correlations indicated by resting-state functional MRI in the (**A**) memory network (MN), (**B**) salience network (SN), (**C**) sensorimotor network (SMN), and (**D**) visual network (VN) with the mean connectivity for individuals with (**i**) low blast exposure and (**ii**) high blast exposure, and (**iii**) the group difference. There were no blast-related differences in (**A**) MN, (**B**) SN, (**C**) SMN, or (**D**) VN functional connectivity as measured by functional MRI. Warm elements in the (**i, ii**) group mean matrices indicate higher connectivity and cool colours indicate lower connectivity; warm elements in the (**iii**) group difference matrices indicate higher connectivity in the high blast group and cool colours indicate lower connectivity in the high blast group.
